# Transdiagnostic Neural Signatures in common Pediatric Psychiatric Disorders: a data-driven meta-analysis of functional neuroimaging studies

**DOI:** 10.1101/2021.03.18.21253910

**Authors:** Jules R. Dugré, Simon B. Eickhoff, Stéphane Potvin

## Abstract

**BACKGROUND:** In the last decades, neuroimaging studies have attempted to unveil the neurobiological markers underlying pediatric psychiatric disorders. However, children diagnosed with such disorders are likely to receive an additional diagnosis in the following years. Yet, the vast majority of neuroimaging studies focus on a single nosological category, which limit our understanding of the shared/specific neural correlates between these disorders. Therefore, we aimed to investigate the transdiagnostic neural signatures through a novel meta-analytical method.

**METHOD:** A data-driven meta-analysis was carried out which grouped similar experiments topographic map together, irrespectively of nosological categories and task-characteristics. Then, activation likelihood estimation meta-analysis was performed on each group of experiments to extract spatially convergent brain regions.

**RESULTS:** One hundred forty-seven experiments were retrieved (3199 subjects): 79 attention-deficit/hyperactivity disorder, 32 conduct/oppositional defiant disorder, 14 anxiety disorders, 22 major depressive disorders. Four significant groups of experiments were observed. Functional characterization suggested that these groups of aberrant brain regions may be implicated internally/externally directed processes, attentional control of affect, somato-motor and visual processes. Furthermore, despite that some differences in rates of studies involving major depressive disorders were noticed, nosological categories were evenly distributed between these four sets of regions. Additionally, main effects of task characteristics were observed.

**CONCLUSIONS:** By using a data-driven meta-analytic method, we observed four significant groups of aberrant brain regions that may reflect transdiagnostic neural signature of pediatric psychiatric disorders. Overall, results of this study underscore the importance of studying pediatric psychiatric disorders simultaneously rather than independently.

## 1. INTRODUCTION

The most prevalent child psychiatric disorders include Attention-deficit/hyperactivity disorder (ADHD), Conduct/Oppositional Defiant Disorder (CD/ODD), anxiety disorders (ANX) and depressive disorders (DEP) and affect approximately 3.4%, 5.7%, 6.5% and 2.6% of children and adolescents in the world, respectively (1). More importantly, evidence suggest that comorbidity between these pediatric psychiatric disorders is the norm rather than the exception. In fact, children with ADHD, CD/ODD, ANX or DEP are likely to being diagnosed with one of these four disorder as a comorbid condition (2-9). Indeed, comorbidity is reported in 51.6-83% of cases, whereas between 3 to 25.8% have received two or more comorbid psychiatric conditions. Although these four diagnostic entities show large comorbidities in children and adolescent, theoretical pathophysiological models that take into account this high level of comorbidity remain largely limited (10).

In the last decade, there has been a growing body of literature suggesting that several genetic (11-15) and environmental risk factors (15-17) which increase the risk for a wide range of psychiatric disorders & psychopathologies may be non-specific. Likewise, recent meta-analyses of structural and functional magnetic resonance imaging studies have shown that adult psychiatric disorders may share several neurobiological deficits (18-22). For instance, during cognitive control tasks, transdiagnostic neural signatures have been identified, which involve the fronto-insular cortex (FIC), the dorsolateral prefrontal cortex and the dorsal anterior cingulate cortex (dACC) to anterior midcingulate/pre-supplementary motor area (aMCC/pre-SMA) and inferior parietal lobule (21). Similarly, adult patients with psychiatric disorders shared prominent deficits in the FIC, amygdala, thalamus and dorso- and ventro-medial PFC during emotion processing tasks (22). Although some differences have been noticed between patients with and without psychotic disorders (21, 22), the search for shared/specific neurobiological features is of great interest for our current understanding of the psychophysiological mechanisms underlying psychiatric disorders.

In neuroimaging literature on childhood/adolescent, studies or meta-analyses that aimed to uncover the specific/transdiagnostic neurobiological markers have been scarce. Indeed, a large majority of task-based fMRI studies has focused on a single psychiatric disorder, therefore limiting our ability to identify common/specific neurobiological markers. Additionally, recent transdiagnostic fMRI meta-analyses have excluded disorders which predominantly emerge in childhood/adolescence such as ADHD and CD/ODD (21, 22). Nevertheless, past meta-analyses and literature reviews on ADHD (23-28), CD/ODD (29-33). ANX (34-39) and DEP (40-47) appear to show qualitatively similar deficits in the anterior insula, medial and lateral prefrontal cortex, amygdala and anterior to midcingulate cortex. Yet, there is a clear need for meta-analytical evidence of transdiagnostic neural signatures in children and adolescents.

Despite the fact that these results may provide substantial insight for our understanding of transdiagnostic neural signatures, classical meta-analytical approaches are prone to important biases. Indeed, authors’ categorization of groups of interest, categorization of fMRI tasks and the choice of task contrast may significantly alter results. In comparison to the classical meta-analytic approach which seeks to identify dysfunctional brain regions in predefined groups of interest, the reverse inference meta-analytical method rather aims to discover groups of interest in a dysfunctional brain region. As such, this novel approach may address the limitations of the classic approach by searching for common/specific neural signatures irrespective of the task-characteristics or the nosological categories. To our knowledge, only one study has investigated transdiagnostic features across adult samples through a reverse-inference meta-analytical method. In fact, the authors observed that when examining diagnosis distribution across brain regions, none of the 56 regions (subcortical and cortical) showed a significant effect of diagnosis across whole-brain studies (48). However, selectively examining diagnosis distribution region after region may yield over-optimistic conclusion about their transdiagnostic characteristics. Given that a single region may be implicated in a wide range of cognitive processes, examining transdiagnostic signatures using a region-of-interest method blurs our ability to capture that co-activation topography may actually differ between psychiatric disorders and reflect functional differences in cognitive processes between disorders. To our knowledge, a meta-analysis aiming to examine transdiagnostic (or specific) groups of regions associated with pediatric psychiatric disorders has never been performed.

Here, we carried out a meta-analysis that primarily aimed to identify groups of aberrant brain regions across psychiatric disorders using a data-driven meta-analytical method. Results from past meta-analyses on adult samples (21, 22) and disorder-specific meta-analyses and literature reviews (23-41) suggest that transdiagnostic features may be expected in FIC (anterior insula/vlPFC), medial and lateral prefrontal as well as in the dorsal anterior and anterior midcingulate cortex. However, considering that deficits in the amygdala is systematically observed in past meta-analyses on adult ANX (35, 36) and DEP (40-47), but less extensively in CD/ODD (29-33) and not found in ADHD (23-28), we hypothesized that the former region would be more closely linked to ANX and DEP than the latter disorders.

## 2. METHODS

### 2.1. Identification of included studies

Our search focused primarily on four diagnostic categories (ADHD, CD/ODD, ANX, DEP) since they all show high comorbidity with each other (2-9). Since meta-analyses and literature reviews on these disorders have been published recently, we extracted data from reference lists of ANX (36, 37, 39), DEP (40, 41, 49), CD/ODD (31), ADHD (28). Inclusion criteria were: (1) original manuscript from a peer-reviewed journal, (2) functional MRI studies that included a fMRI task, (3) use of a whole-brain methodology (i.e., studies using region-of-interest were excluded), (4) < 18 years old participants meeting criteria for at least one of the following pediatric psychiatric disorder: (a) Attention deficit with/or without hyperactivity; (b) Disruptive disorder (Conduct disorder and/or Oppositional Defiant Disorder); (c) Anxiety disorders (i.e., Posttraumatic Stress Disorder, Generalized Anxiety Disorder, Social Anxiety Disorder) and/or (d) Unipolar Major Depressive Disorder.

### 2.2. ALE Method

ALE approach was used in the current coordinate-based meta-analysis in order to extract spatially convergent peaks across pediatric psychiatric disorders (GingerALE version 3.0.2, http://www.brainmap.org/ale/). Study results were extracted, irrespectively of the direction (decreased/increased) of the diagnosis or task-contrast effect, to create an aberrant activation map. Two experiments from the same study were considered as distinct if they included two different samples or two different fMRI tasks. Each experiment’ pooled task-contrasts was manually annoted and categorized if they included: a cognitive component (i.e., response inhibition, attentional processes); an emotional component (i.e., response to positive stimuli; to negative stimuli; to both). These categorizations were not mutually exclusive (e.g., emotional go-no/go or Stroop). Rates of boys in each sample and percentage of subjects that received medication were also extracted for each experiment. Coordinates of experiments that were reported originally in Talairach stereotaxic space were converted into MNI (Montreal Neurologic Institute) space before using them in the analyses.

First, a modeled activation map (MA) was created by modeling coordinate foci (x,y,z) with a spherical Gaussian probability distribution, weighted by the number of subjects in each experiment. This is performed to account for spatial uncertainty due to template and between-subject variance (50), and ensure that multiple coordinates from a single experiment does not jointly influence the modeled activation value of a single voxel. Voxel-wise ALE scores were then computed as the union of MA maps, which provide a quantitative assessment of convergence between brain activation across experiments. Then, these maps were cut off by a cluster-forming threshold. In fact, the size of the supra-threshold clusters was compared against a null distribution of cluster sizes derived from artificially created datasets in which foci were shuffled across experiments, but the other properties of original experiments (e.g., number of foci, uncertainty) were kept. Finally, this resulted in calculating the above chance of observing a cluster of the given size (51). In the current study, we use the following statistical threshold: a voxel-level cluster forming threshold of p<0.001 and a cluster-level family-wise correction (pFWE < 0.05), with 5,000 permutations (52).

We performed a disorder-specific meta-analysis using these “*classical*” steps of the ALE method for each diagnostic group, separately.

### 2.4. Neurobiologically-driven Meta-analytical procedure

#### 2.4.1. Modeled Activation & Cross-Correlation Matrix (Step 1 & 2)

Modeled activation (MA) map was created for each experiment (2mm^3^ resolution) (Figure 1, Step 1). Each resulting MA map was converted into a 1D feature vector of voxel values (i.e., 2mm^3^ grey matter mask in MNI space) and concatenated together to form an experiment (*e*) by voxel matrix (*v*) (147 experiments x 226,654 voxels). Pairwise Spearman’s rank correlation was performed between the 1D feature vector of each experiments to obtain spatial similarity between maps (*e* by *e* symmetric correlation matrix) (Figure 1, Step 2).

**Figure 1.**
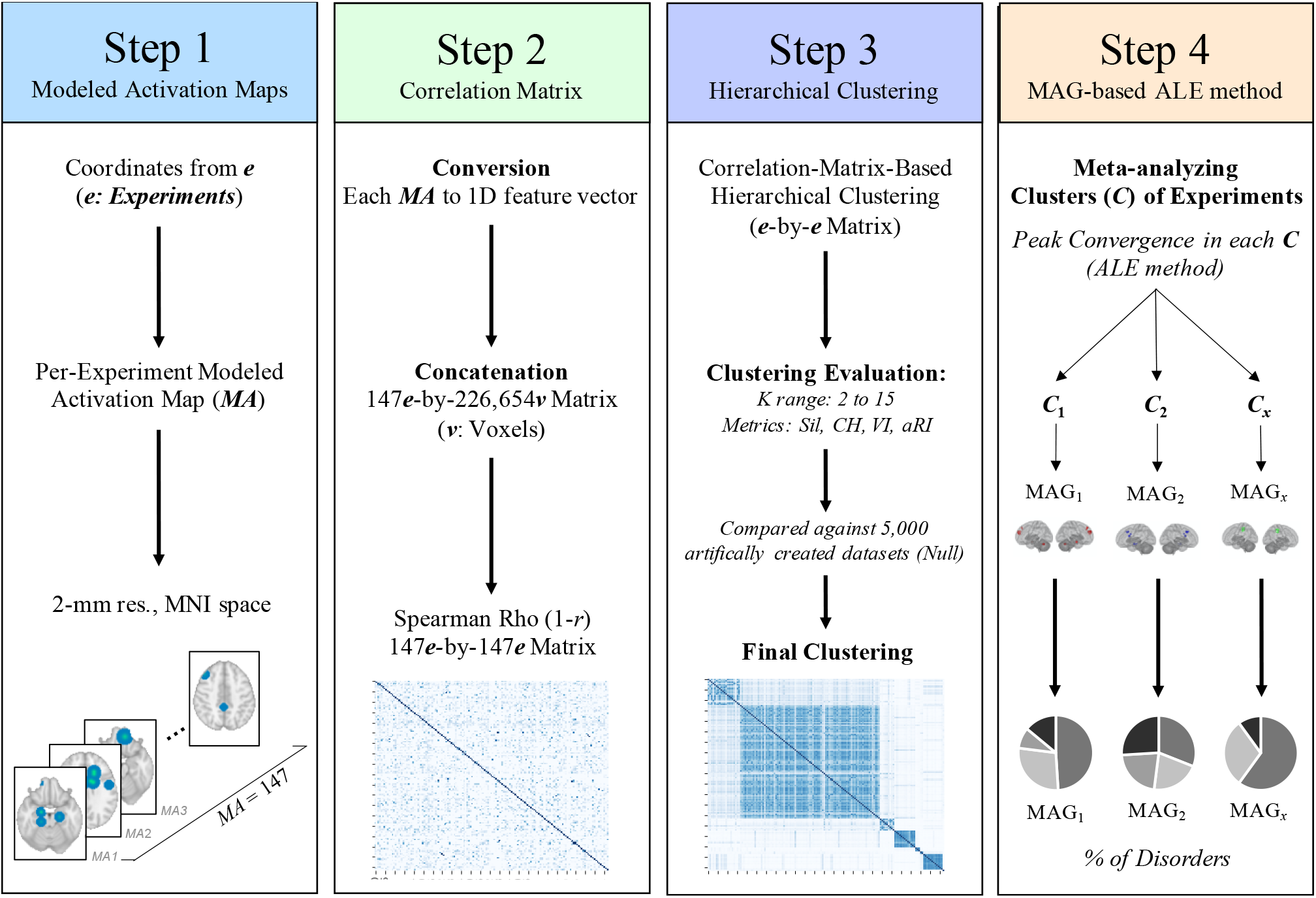
Workflow of the current study. **Step 1:** Creation of a MA map for each experiment, weighted by sample size. **Step 2:** Pairwise Spearman Rho correlation was performed between every MA map. **Step 3:** Clustering analysis was performed on the correlation matrix to extract groups of experiments sharing similar MA map. **Step 4:** ALE meta-analysis was conducted on experiments within each group. Phenotype assessment was then carried out to investigate under/over-representativeness of disorders, sample and task characteristics across identified groups.

#### 2.4.2. Correlation-Matrix-Based Hierarchical Clustering (Step 3)

In order to extract data-driven groups of experiments that showed similar brain topographic map, we performed a Correlation-Matrix-Based Hierarchical Clustering (CMHC) analysis, as previously used on meta-analytic data (53, 54). The CMHC was carried out using correlation distance (1 – *r*) (Figure 1. Step 2) and average linkage method. We thus repeated the CMHC for a range of 2 to 15 MAGs. The optimal number of MAGs was determined by comparing frequently used metrics in fMRI clustering (i.e., silhouette and calinski-harabasz indices, variation of information & adjusted rand index) (55), for each of the 2-15 MAGs (See Supplementary Material for more information). More precisely, we used a 90% subsampling resample strategy (without replacement, 5000 iterations). This implied that we randomly removed 10% of experiments, iteratively, to perturbate the ability to group the most correlated experiments with each other, successively. Then, for K range, each metric was compared against a null distribution. To do so, 5,000 datasets were created artificially by shuffling foci locations across experiments but preserving original experiments’ properties (e.g., number of foci, sample size). The average of each metric (i.e., silhouette and calinski-harabasz indices) derived from *true* dataset, was normalized using this null distribution (Average_TRUE_ – Average_NULL_ / STD_NULL_) and then plotted for K range (2-15 MAGs). This improves our ability to select the optimal K by taking into account the probabilities of getting a certain metric value in a random spatial arrangement. Given that the ground truth class labels are unknown, we compared, for each K, the consistency (adjusted rand index) and shared information distance (variation of information) between Label_TRUE_ with the Label_NULL_ then averaged across the 5000 iterations. These metrics were then plotted for K range. A local minimum in the plot suggests a decrease in overlap between both sets of labels.

After finding the optimal number of MAGs, the most stable label solution was found by grouping experiments that were labelled similarly across the 5,000 subsampling iterations. More precisely, we calculated the hamming distance between each experiment’s label ids (147 experiments by 5000 iterations ids) to calculate the proportion of disagreement between two experiments’ set of labels. A CMHC was performed on a final 147-by-147 experiments’ distance matrix, representing the distance between each pair of experiments. We then used the determined most optimal number of MAGs to extract the most stable label ids.

Finally. MAGs with less than 10 experiments were considered as outliers and excluded from further analyses, since analyses involving < 10 experiments drastically increases the risk that a single experiment drives the results (52). All these analyses were performed using Scikit-learn (version 0.21.3) in Python (version 3.7.4) (56).

#### 2.4.3. Meta-Analytical Groupings (Step 4)

Experiments (*e*) within each meta-analytical grouping (MAG) were then meta-analytically processed (Step 4), using the activation likelihood estimate (ALE) algorithm (GingerALE version 3.0.2) (50, 51). This was performed to extract significant peaks convergence across each MAG (derived from Step 3). To examine under- and overrepresentations of nosological categories, task and sample characteristics within each MAG, we carried out one-tailed binomial tests comparing their prevalence with their base rate (across all experiments). Main effects of diagnosis, task and sample characteristics between MAGs were investigated through chi-squares (*X*^2^) and Kruskal-Wallis (H) tests. Literature bias was also assessed to compare differences between nosological categories in terms of task and sample characteristics (See Supplementary Material).

Finally, for each MAG, we extracted functional characterization using the *Behavioral Analysis plugin* of the Multi-Image Analysis GUI (57). A Z-score was calculated using binomial tests for 83 paradigm classes and 51 behavioral subdomains across more than 10,000 experiments. A z-score higher or equal to 3 is considered significant (i.e., equaling p<0.05 Bonferroni corrected for multiple comparisons).

## 3. RESULTS

### 3.1. Identified studies and characteristics

A total of 124 original studies met the inclusion criteria for the meta-analysis, of which 11 involved more than one sample and 8 comprised two or more distinct fMRI task contrasts. This resulted in 147 experiments (1030 foci) involving 3199 cases that were compared to 3024 healthy controls. Also, mean age of cases was 13.8 years old (SD=2.25) and the average rate of boys across samples was 71.67%. (see Supplementary Table for complete list of studies and their respective characteristics). Interestingly, disorder-specific studies showed significant literature bias regarding the choice of neurocognitive task domains, average of sex ratio, and the average of prescribed medication per samples (See Supplementary Table).

### 3.2. Classical ALE Meta-analyses per-disorder category

Meta-analyses were performed across all foci, independently of the activation direction, for ANX (46 foci, 14 experiments, 293 subjects), DEP (103 foci, 22 experiments, 388 subjects), ADHD (700 foci, 79 experiments, 1892 subjects) and CD (197 foci, 32 experiments, 626 subjects). However, each disorder-specific meta-analysis revealed no significant spatial convergence (See Supplementary Material for results using uncorrected threshold). Nonetheless, we observed peak convergence in dorsal/perigenual ACC (MNI x=-8, y=38, z=8, maximum ALE score = 0.0167, 129 voxels) for the internalizing category (ANX+DEP) and pre-SMA (x=4, y=26, z=44, maximum ALE score=0.0279, 147 voxels) for the externalizing category (ADHD+CD/ODD). Additionally, merging experiments across pediatric psychiatric disorders, we observed a significant cluster that included voxels of the right anterior MCC and the pre-supplementary motor area (x=4, y=24, z=42, ALE score=0.021, 323 voxels). Despite the fact that ADHD (57.14%) and CD/ODD (35.71%) largely contributed to this specific brain region, proportions did not significantly differ from their respective base rate. Furthermore, no significant differences were observed regarding nosological categories nor task-characteristics. However, Mann-Whitney U revealed a significant sex effect, indicating that the aMCC/pre-SMA may be associated with higher average rates of boys across samples (M=69.7% versus M=91.77%; U=479.0; p=0.003). No other significant differences were observed.

### 3.3. Neurobiologically-driven meta-analysis

#### 3.3.1. Clustering Solution

Clustering solutions were investigated for a range of *K* = 2-15 MAGs with resampling method (90% subsamples and 5,000 iterations). Average of the 5,000 iterations metric values for each *K* were plotted. Despite that the Calinski-Harabasz index exhibited a monotonic behavior (constantly increasing), the silhouette index showed a stable solution at K=8. Furthermore, aRI plot showed lowest scores at K=3 and K=8, while the greatest increases in variation of information were observed moving from K=2 to K=3, from K=6 to K=7 and K=7 to K=8. Based on these results, the solution with 8 MAGs was found as the most optimal (See Supplementary Figure 1).

Of the 8 MAGs, 4 comprised less than 10 experiments (n=8, 3, 2 & 1, respectively). These were excluded from further analyses. The remaining 4 MAGs represented 90.58% of total sample of experiments (133 experiments out of 147): MAG1 (577 subjects, 21 experiments and 120 foci), MAG2 (1848 subjects, 87 experiments, 708 foci), MAG3 (197 subjects, 13 experiments, 52 foci), MAG4 (278 subjects, 12 experiments, 113 foci) (Figure 2).

**Figure 2.**
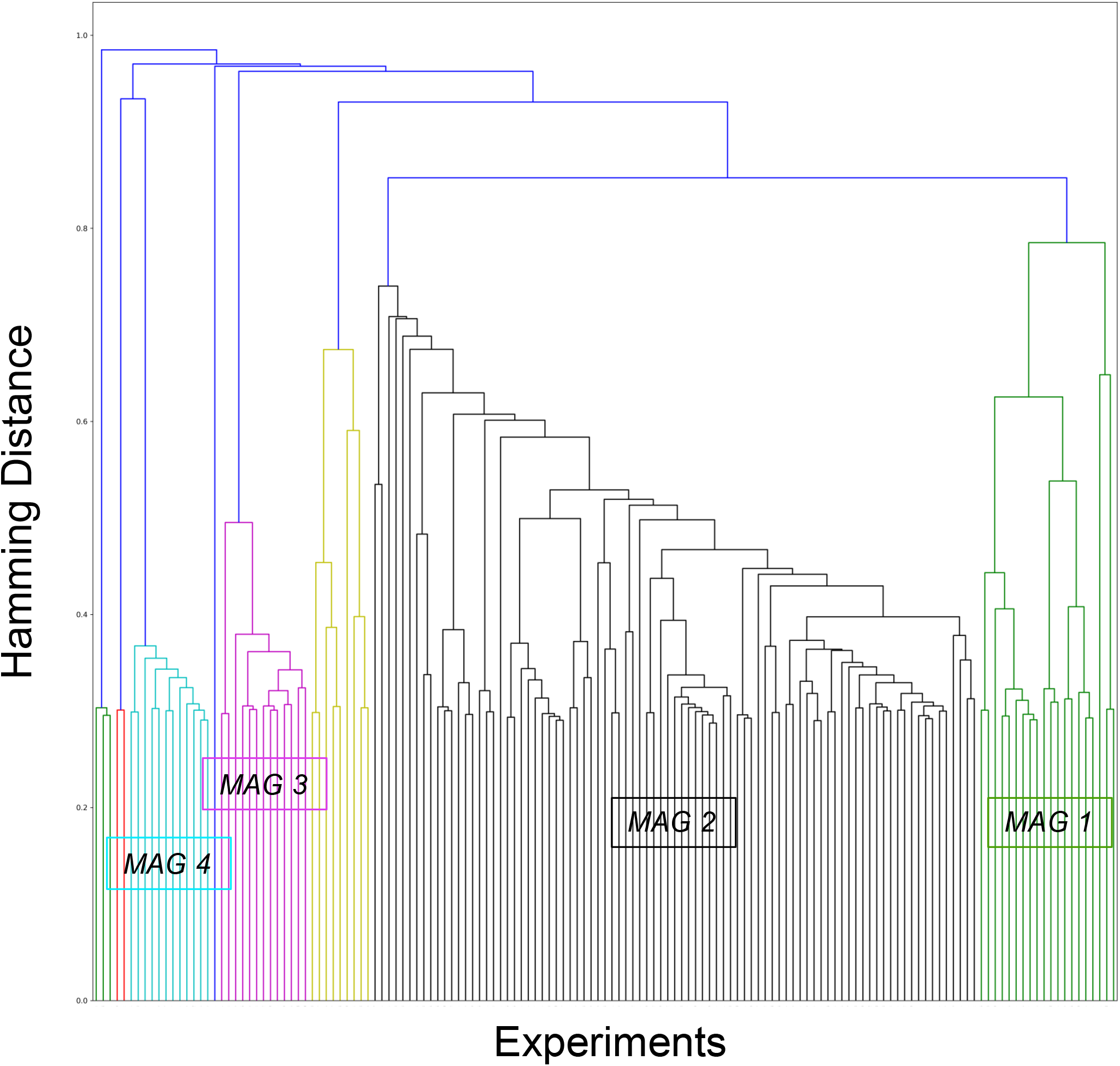
Hierarchical clustering of aberrant activation maps. This dendrogram represents the final hierarchical clustering model which grouped experiment showing similar aberrant activation maps. The 4 significant meta-analytical groupings (MAGs) represented 90.58% of total sample of experiments: MAG1 (green) = 21 experiments and 577 subjects; MAG2 (black) = 87 experiments (1848 subjects); MAG3 = 13 experiments (197 subjects) & MAG4 (cyan) = 12 experiments (278 subjects).

#### 3.3.2. ALE Meta-analysis

ALE meta-analysis was performed across experiments in each of the four MAGs, separately, to examine spatial convergence across similar experiments (p<0.001 voxel-level, p<0.05 cluster-level FWE-corrected). As shown in Table 1 and Figure 3, experiments of the MAG-1 had convergent peaks in the right rostrodorsal dorsomedial PFC (dmPFC) and the left caudal dmPFC (see (58)), the left cerebellum (Lobule VI), the right dorsolateral prefrontal cortex (dlPFC, see Cluster 5, Brodmann area (BA) 9/46d, (59)) and the middle temporal gyrus (MTG). The ALE meta-analysis for MAG2 included the right anterior MCC (BA32’, see (60, 61)) and the pre-supplementary motor area, the left amygdala (encompassing the hippocampus and parahippocampus) and the left aMCC (BA24 a’-b’, see (60, 61)). Regarding the MAG3, significant aberrant map revealed spatial convergence in the right posterior precentral (BA4p) to postcentral gyri (BA2-3), the right supramarginal gyrus, inferior parietal lobule, human IntraParietal area 2 and the left postcentral gyrus (BA2) and the inferior parietal lobule (PFt area, see (62)). Finally, spatial map of MAG-4 included occipital/cerebellar regions such as bilateral ventral extrastriate cortex (right area hOc3v-hOc4v and left hOc4v, (63)), bilateral fusiform gyrus (Area FG4), bilateral Lobule VI, left calcarine gyrus (Area hOc1) as well as right posterior middle/inferior temporal gyrus (hOc1).

**Table 1.** **ALE** meta-analysis results of each significant groups of experiments

| MAGs | Clusters | Size (mm<sup>3</sup>) | MNI |  |  | ALE | Cluster Breakdown |
| --- | --- | --- | --- | --- | --- | --- | --- |
|  |  |  | Coordinates |  |  |  |  |
|  |  |  | X | Y | Z |  |  |
| MAG1 | 1 | 2456 | 14 | 46 | 28 | 0.0175 | R dmPFC (rostrrodorsal) |
|  | 2 | 1152 | -16 | 56 | 22 | 0.0154 | L dmPFC (caudal) |
|  | 3 | 1096 | -24 | -60 | -28 | 0.0177 | L Cerebellum (Lobule VI) |
|  | 4 | 1048 | 30 | 40 | 46 | 0.0164 | R dlPFC |
|  | 5 | 848 | 58 | -8 | -18 | 0.0216 | R MTG/STG |
| MAG2 | 1 | 1352 | 8 | 18 | 40 | 0.0243 | R aMCC (Area 32')/pre-SMA |
|  | 2 | 1296 | -20 | -10 | -16 | 0.0272 | L Amygdala |
|  | 3 | 1040 | -2 | 12 | 22 | 0.0321 | L dACC (Area 24a'-b') |
| MAG3 | 1 | 976 | 34 | -26 | 52 | 0.0125 | R Pre-/Postcentral gyri (Area 2-3 & 4p) |
|  | 2 | 800 | 46 | -34 | 44 | 0.0133 | R Supramarginal gyrus (Area 2, Pft) |
|  | 3 | 720 | -42 | -32 | 42 | 0.0112 | L Postcentral gyrus (Area 2, Pft) |
| MAG4 | 1 | 2336 | 20 | -78 | -12 | 0.0172 | R Lingual (h0c3v) |
|  | 2 | 1224 | -18 | -66 | -24 | 0.0159 | L Cerebellum (Lobule V1) |
|  | 3 | 968 | 44 | -58 | -4 | 0.0168 | R pMTG |
|  | 4 | 912 | -44 | -48 | -14 | 0.0151 | L pITG |
|  | 5 | 736 | -8 | -62 | 6 | 0.0186 | L Calcarine Cortex |
|  | 6 | 728 | 40 | -52 | -26 | 0.0155 | R Cerebellum (Lobule VI) |

Note. MAG = Meta-analytical Grouping; PFC = Prefrontal Cortex; dmPFC = dorsomedial PFC; dlPFC = dorsolateral PFC; MTG = Middle Temporal Gyrus; STG = Superior Temporal Gyrus; aMCC = anterior MidCingulate Cortex; pre-SMA = pre-supplementary motor area; dACC = dorsal anterior cingulate cortex; IPL = Inferior Parietal Lobule; SPL = Superior Parietal Lobule; pMTG = posterior MTG; pITG = posterior ITG.

**Figure 3.**
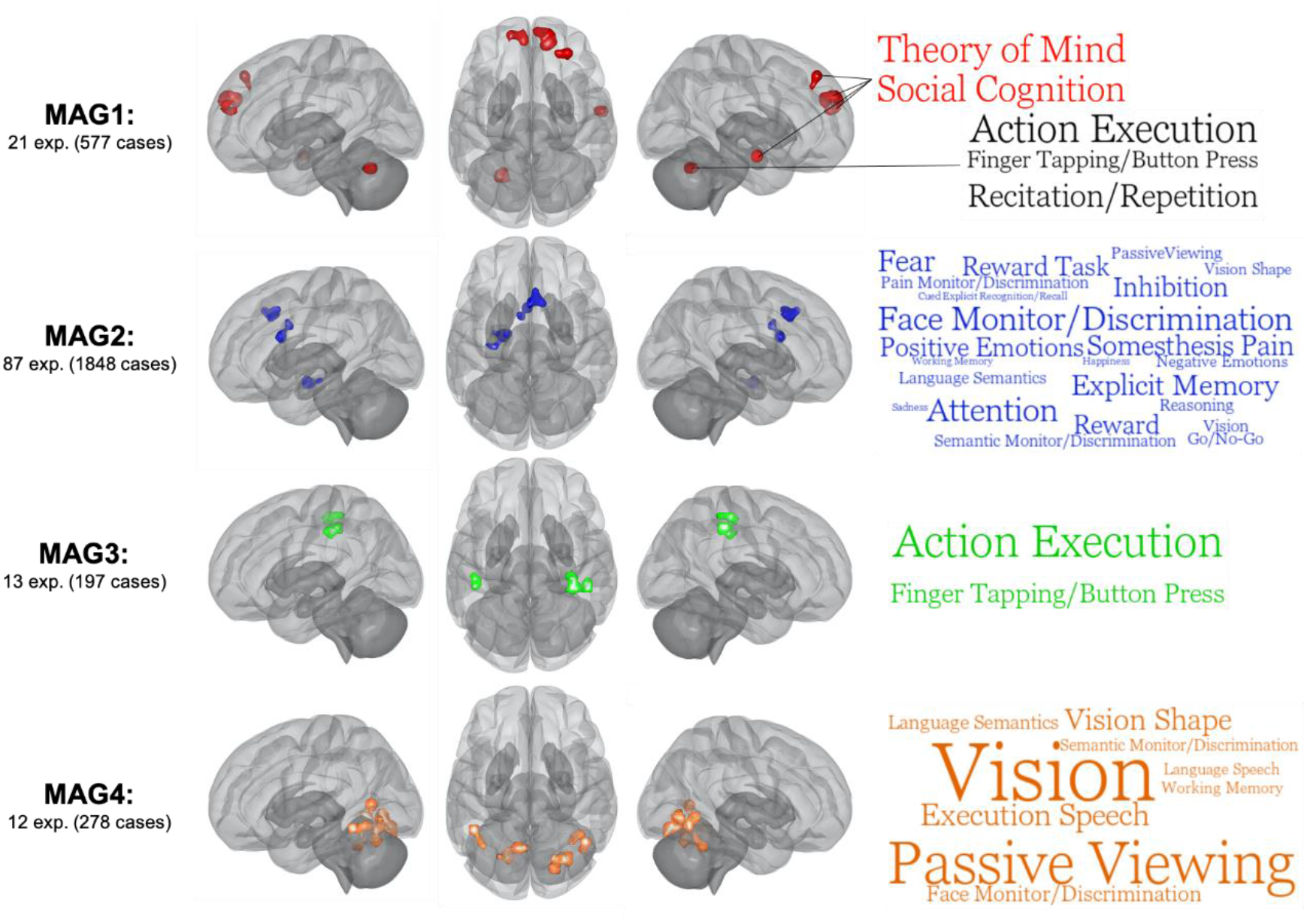
ALE meta-analysis on each significant meta-analytical grouping (MAGs). Images are shown for left hemisphere (lateral), superior view and right hemisphere (lateral) respectively. ALE images were thresholded at p<0.001 at the voxel-level and pFWE > 0.05. Word clouds were generated using BrainMap database terms (Behavioral Subdomains & Paradigm). Font size represents Z-score associated with the whole MAG (all words are significant p=0.05 with Bonferroni correction).

#### 3.3.3. Functional characterization of MAGs

Functional characterization of MAGs (i.e., MAG-wide & cluster-specific) was performed to examine their relationships with behavioral domains and paradigms of the BrainMap database (see Figure 3, Supplementary Material):

##### MAG1

Experiments mainly included response inhibition (7) and reward decision-making tasks (5, e.g., monetary incentive delay task, Colorado balloon game). Functional characterization using the BrainMap database yielded no significant behavioral/paradigm classes. However, bilateral dmPFC and anterior MTG/STG were positively associated (Z>3.0) with social cognition/theory of mind, and negative related (Z<-3.0) with action execution. Interestingly the left Lobule VI show positive association with action execution and negative relationship with social cognition, whereas dlPFC was related to action inhibition. In sum, this MAG may be characterized by deficits of co-occurrent brain regions subserving social cognition during cognitive & reward decision-making tasks.

##### MAG2

Experiments within this MAG primarily included task contrasts comprising an emotional component (k=42) of which 24 used negative emotional stimuli (e.g., negative emotional faces). Other main task domains were attentional, cognitive control and reward tasks. MAG2 was characterized by a wide range of behavioral subdomains and paradigms from the BrainMap Database including attention, face monitoring & discrimination and explicit episodic memory. Furthermore, the right aMCC/pre-SMA (Attention) shared similar cognitive domains with left amygdala (Face Monitoring/Discrimination) such as explicit memory, semantic monitoring and positive emotions/reward. Also, the right aMCC/pre-SMA and the left dACC (Somesthesis pain) were both associated with pain monitoring & discrimination paradigm and somesthesis pain domain. However, the left amygdala and the left dACC did not share any behavioral subdomains or paradigms. Thus, given these findings, the co-occurrence of the dACC, aMCC/pre-SMA and the amygdala may be implicated in stimulus-driven attentional control.

##### MAG3

Experiments within this MAG included a variety of cognitive and sensorimotor tasks (e.g., finger sequencing, anti-saccade, mental rotation, nback). Using the BrainMap Database, we observed that MAG3 was significantly associated with action execution and finger tapping/button press. Region-specific analyses revealed that the three regions, the right posterior precentral/postcentral (Action Execution), the right SMG (Action Execution) and left postcentral (Action Execution), shared action execution, finger tapping and somesthesis behavioral domains. Additionally, only the right precentral and the left postcentral clusters were both associated with tactile monitoring. In sum, brain regions of this MAG may encompass sensorimotor/action execution processes.

##### MAG4

Finally, experiments from the MAG4 mainly included various cognitive tasks (10). Functional characterization using the BrainMap database revealed significant associations with vision, passive viewing and speech execution. Region-specific analyses revealed that all but the calcarine were related to vision. Furthermore, the right pMTG/ITG, the left pITG/FF and the right lobule VI shared face monitoring/discrimination, passive viewing, vision shape and covert naming domains. In short, MAG4 may reflect co-occurrent deficits in brain regions involved in visual processing during cognitive tasks.

#### 3.3.4. Phenotype Assessment 1: Nosological Categories

MAG1 was less likely to include DEP samples (X_2_ = 4.16, p=0.041), compared to all the other MAGs. Indeed, proportions of DEP samples in MAG1 was significantly lower than its base rate (0% versus 15.00%, one-tailed p=0.028) (see Table 2). Considering the between-disorder literature bias revealed that the lower rates of DEP samples in MAG1 were replicated when restricting experiments to those using an emotional task contrast and mixed sex samples (Supplementary Material)

**Table 2.** Characteristics of Experiments across meta-analytical groupings

| Characteristics | Total (n=147) |  | MAG1<br>(k=21) |  | MAG2<br>(k=87) |  | MAG3<br>(k=13) |  | MAG4<br>(k=12) |  |
| --- | --- | --- | --- | --- | --- | --- | --- | --- | --- | --- |
|  | n | % | n | % | n | % | n | % | n | % |
| *<u>Nosological Categories</u>* |  |  |  |  |  |  |  |  |  |  |
| ADHD | 79 | 53.7% | 14 | 66.7% | 43 | 49.4% | 8 | 61.5% | 8 | 66.7% |
| CD | 32 | 21.8% | 4 | 19.0% | 17 | 19.5% | 4 | 30.8% | 3 | 25.0% |
| ANX | 14 | 9.5% | 3 | 14.3% | 9 | 10.3% | 0 | 0.0% | 1 | 8.3% |
| DEP | 22 | 15.0% | 0*† | 0.0% | 18† | 20.7% | 1 | 7.7% | 0 | 0.0% |
| *<u>Task-contrast Domain</u>* |  |  |  |  |  |  |  |  |  |  |
| Cognitive | 88 | 59.9% | 10 | 47.6% | 53 | 60.9% | 9 | 69.2% | 10 | 83.3% |
| Response Inhibition | 44 | 29.9% | 7 | 33.3% | 29 | 33.3% | 3 | 23.1% | 4 | 33.3% |
| Attention | 23 | 15.6% | 1 | 4.8% | 13 | 14.9% | 3 | 23.1% | 3 | 25.0% |
| Emotion | 71 | 48.3% | 12 | 57.1% | 42 | 48.3% | 3* | 23.1% | 5 | 41.7% |
| Positive | 17 | 11.6% | 6*† | 28.6% | 7* | 8.0% | 0 | 0.0% | 1 | 8.3% |
| Negative | 37 | 25.2% | 4 | 19.0% | 24 | 27.6% | 1 | 7.7% | 2 | 16.7% |
| Both | 16 | 10.9% | 2 | 9.5% | 10 | 11.5% | 2 | 15.4% | 2 | 16.7% |
| *<u>Sample Characteristics</u>* |  |  |  |  |  |  |  |  |  |  |
| Medication-Naïve | 61 | 41.5% | 14† | 66.7% | 40 | 46.0% | 6 | 46.2% | 5 | 41.7% |
| Average Med per sample | - | 26.7% | - | 35.2% | - | 26.3% | - | 19.4% | - | 20.9% |
| Mixed Sex Sample | 95 | 64.6% | 12 | 57.1% | 60 | 69.0% | 7 | 53.8% | 8 | 66.7% |
| Average Boys per Sample | - | 71.7% | - | 77.6% | - | 71.4% | - | 76.1% | - | 61.1% |

Note. \* represent significant difference compared to its base rate (one-tailed $p < 0.05$ ). † represents significant differences between MAGs ( $p < 0.05$ )

Additionally, MAG2 had more DEP samples than other MAGs (X_2_ = 8.43, p=0.004). However, compared to its base rate, proportion of DEP samples was not significantly overrepresented in MAG2 (20.7% versus 15.0%, one-tailed p=0.123) (see Table 2). After considering the between-disorder literature bias, we observed that the higher rates of DEP samples in MAG2 was replicated when restricting experiments to those using a cognitive task contrast or an emotional task contrast but also in experiments with only medication naïve sample and in mixed sex samples (Supplementary Material)

#### 3.3.5. Phenotype Assessment 2: Task & Sample Characteristics

First, we observed that the rate of experiments within the MAG1 that included a positive emotional stimulus was higher than other MAGs (X_2_ = 8.62, p=0.003), and significantly overrepresented compared to its base rate (28.6% versus 11.6%, one-tailed p=0.028) (see Table 2).

Additionally, experiments in MAG2 were less likely to include positive emotional task contrast (X_2_ = 3.97, p=0.046) and marginally associated with greater experiments with negative emotion task contrast (X_2_ = 3.31, p=0.069) (see Table 2), compared to other MAGs. However, proportions of these task domains were not statistically different than their base rates.

MAG3 had significantly lower rate of general emotional stimuli compared to other MAGs (X_2_ = 4.20, p=0.040), which was marginally lower compared to its base rate (37.5% versus 48.3%, one-tailed p=0.059). Other characteristics did not reach statistical significance, compared to their base rates.

Also, despite that the MAG1 had significantly higher rate of medication-naïve subjects, compared to its base rate (one-tailed p=0.018), MAGs did not differ in rates of experiments with medication-naïve samples (X^2^ = 2.25, p=0.522), in the average rate of prescribed medication (Kruskal-Wall H=2.74, p=0.433). No differences were observed concerning the rate of mixed sex samples (X^2^ =1.90, p=0.594) and in the average rate of boys in samples (Kruskal-Wall H=2.40, p=0.493).

## 4. Discussion

The current meta-analysis was carried out to examine the shared and/or specific neural correlates of pediatric psychiatric disorders (ADHD, CD/ODD, ANX & DEP). To do so, we used a novel data-driven meta-analytical method that aimed to extract groups of experiments which show similar brain topographic maps. We identified 4 significant MAGs, which comprised co-occurrent deficits in brain regions that may share features with (1) internally/externally directed processes; (2) attentional control of emotions, (3) action execution and (4) visual processes. More importantly, we found underrepresentation of DEP samples in MAG1 but overrepresentation of samples in MAG2. No other significant differences in nosological categories between MAGs we found.

MAG1 included bilateral dmPFC, dlPFC, MTG/STG and Lobule VI. More precisely, we observed that dmPFC-MTG/STG were involved in social cognitions, whereas dlPFC and Lobule VI were characterized as action inhibition and execution, respectively. We also found main task-effect of the utilization of a positive emotional stimulus. Interestingly, several findings suggest that during some cognitively demanding tasks, brain regions involved in internally directed processes (e.g., dmPFC & anterior MTG/STG) flexibly shifts their activity to enable goal-directed processes (e.g., dlPFC & Lobule VI) (64-66). Given these data, deficits in brain regions involved in MAG1 may reflect a failure to disengage internal processes at the cost of goal-directed processes (67). Interestingly, our results suggest that these co-occurrent deficits (i.e., dmPFC, dlPFC, Lobule VI and MTG/STG) are less likely to be reported in that DEP samples given that none of the DEP samples was grouped in MAG1. However, a recent study has shown lower thickness in regions spanning the MAG1 (i.e., encompassing the fronto-parietal and default-mode network) associated externalizing but also internalizing factors (68). Furthermore, meta-analytical evidence suggests dysconnectivity between the default-mode and dorsolateral prefrontal cortex (fronto-parietal network) in DEP (69). Given that these findings were observed in adult samples and that anticorrelation between these processes varies from childhood from adulthood (70), it is possible that these deficits may rather be observed in adulthood DEP. Additionally, DEP samples did not frequently use positive emotional stimuli (k=3 out of 22), which was found to be main task-characteristic of MAG1. As such, this may suggest that the lack of relationship between MAG and DEP may be explained by task differences. Considering the small sample size involved, we cannot completely rule out the possibility of such deficits in children with DEP.

The largest MAG (MAG2, k=87) was constituted of the aMCC/pre-SMA, amygdala and dACC. This MAG was mainly characterized by attention, face monitoring and explicit episodic memory, using the BrainMap database. Interestingly, deficits in these brain regions were associated with higher rates of DEP samples, in comparison with other MAGs. This is somewhat expected given the large number of meta-analytical evidence consistently showing aberrant activation in these particular regions during negative emotional tasks in adult with major depression (42-47). However, no effect was observed in ANX samples, potentially due to the limited sample size. Nonetheless, deficits in these regions were also observed across adult ANX & DEP samples, during negative emotion processing (20, 22, 71). In addition, we observed a marginally significant association between this MAG and negative emotional stimuli, indicating a possible task-effect. In fact, although rates of ADHD (≈50%), CD/ODD (≈20%) did not differ between other MAGs, evidence suggests that these disorders may also show deficits in MAG2 regions, particularly during emotion processing tasks (31, 72) which correlates with general psychopathology score (68, 73, 74). In sum, this MAG may reflect general deficits in emotional lability, inherent to DEP, yet frequently observed in children/adolescent with ADHD (75) and/or CD (76).

We also found deficits in brain regions (e.g., pre- and postcentral gyrus) subserving action execution/finger tapping tasks (MAG3). This MAG was less likely to comprise emotional tasks, which is consistent with the fact that emotional tasks usually require less motor execution. Interestingly, deficits in similar regions (i.e., somato-motor network) were also observed in a recent study showing significant transdiagnostic association with general maladaptive functionality (77). Although deficits in these regions are currently not well understood, sensory deficits such as tactile perception and body awareness are often reported in children with pediatric psychiatric disorders (78-82). It is thus possible that abnormalities in MAG3 may reflect deficits in tactile perception, crucial for accurate performance of purposeful movements (83) such as in cognitive tasks.

Finally, we found evidence of early processing deficits across disorders (MAG4). However, this MAG was not specifically associated with sample- or task-characteristics. Recent studies have shown replicable structural alterations in brain regions spanning this MAG. In fact, the authors demonstrated, through two different samples comprising 1246 (84) and 875 (85) subjects, that the general psychopathology factor score was associated with deficits in occipital (i.e., Lingual, Calcarine cortex) and cerebellum regions. These regions are implicated in variety of visual functions such as detecting relevant changes in the environment (e.g., visual oddball) (86, 87). Thus, MAG4 may mirror several dysfunctional processes in early visual processing, including gazing at task-irrelevant stimuli. For example, during face-emotion tasks, the number and duration of fixation to the eye regions have been reported to be significantly lower in ADHD with and without CD (88), in childhood psychopathic traits (89), in ODD/CD (90-92), anxiety disorders (93-95) (96)) and depression (97, 98). Likewise, deficits in the ability to filter out irrelevant stimuli are also observed in continuous performance test (99) and visual search tasks (98) in these populations.

In the classical transdiagnostic approach, we observed significant overlaps in the aMCC/pre-SMA. Furthermore, we found that externalizing disorders (i.e., ADHD, CD/ODD) were associated with deficits in the pre-SMA, whereas internalizing disorders (i.e., ANX, DEP) yielded aberrant activity in the dorsal/perigenual ACC. These concur with recent transdiagnostic findings in adult samples showing aberrant activation during cognitive control tasks (dACC & aMCC (21)). It is worth mentioning that in children diagnosed with a psychiatric disorder, more than half will receive, *at least*, an additional diagnosis than their primary one in the following years (100-105). Also, comorbidity rates are higher within externalizing (between CD/ODD & ADHD) and internalizing (between DEP and ANX) disorders than between these broad categories. However, it remains unknown whether this high comorbidity rate in children is due to a common vulnerability (e.g., shared risk factors) or the presence of cross-cutting criteria (e.g., impulsivity, neuroticism). Hence, this issue unequivocally needs to be tackled in the future. In addition, we observed no significant peak convergence across each of the disorder-specific meta-analysis. Indeed, this lack of convergence concurs with results from recent meta-analyses which revealed similar results in CD/ODD, ADHD and DEP, using a somewhat conservative threshold (p<0.001, cFWE<0.05) (28, 31, 106). These null findings were also observed even when examining specific neurocognitive task domains. Despite that this lack of convergence might have been attributable to a number of between-study differences (e.g., stimulus, sex effect, statistical threshold, sample size), one possibility that deserves careful attention is the within-disorder heterogeneity. Indeed, it is generally well accepted that DSM-derived categories comprise subfactors that are characterized by different psychological processes (107-112). Thus, this plurality in criteria substantially increases the risk of finding distinct set of symptoms while still meeting the diagnostic threshold (from 42 [GAD] to 116,200 [ADHD] theoretical set of criteria, (113, 114). Therefore, we could not rule out the possibility that increasing the sample size in meta-analyses, which also increase the between-sample heterogeneity, may reduce the ability to detect robust findings. In sum, irrespectively of the nosological categories or task-characteristics, we found 4 significant groups of aberrant brain regions in children/adolescent with a psychiatric disorder, which comprised relatively similar rates of ADHD, ANX, CD/ODD (and DEP to a lesser extent).

## LIMITATIONS

First, we performed cluster analysis across pediatric psychiatric disorders and across fMRI paradigms. Since there were limited data available to perform domain-specific analyses, it is possible that our results may have been altered by literature bias (see Supplementary Material) concerning the use of particular neurocognitive task domains per diagnosis category. However, subanalyses were carried out to examine these confound effects. In the following years, as more samples using different tasks domains will be available, more precise analyses will be possible. Second, the limited sample size for the ANX samples (k=14) may have explained the null findings in *classical* disorder-specific meta-analysis as well as the lack of over/underrepresentation across MAGs. It has been recommended that a sample size greater than 17 experiments are required to achieve good statistical power in *classical* meta-analyses. Finally, we used hierarchical clustering with spearman correlation as distance measure and average linkage algorithm. Although these parameters are frequently utilized in studies using similar meta-analytical approaches, it is possible that the most optimal set of parameters would have been specific to our study.

## CONCLUSIONS

In sum, we observed transdiagnostic neural signatures across common pediatric psychiatric disorders. More particularly, the identified groups of co-occurrent deficits spanned brain regions that share features with internally/externally directed processes, emotional lability, somato-motor & visual processes. We also observed that DEP samples were less likely to display aberrant co-activation map involving internally/externally directed processes, but more likely to exhibit deficits in brain regions implicated in attentional control of emotions. Also, these MAGs did not specifically fit particular neurocognitive domains. Indeed, they rather involved multiple subprocesses (e.g., Self-reflective & Execution/Inhibition; Threat system & Attentional Control). Thus, future studies may benefit from examining the task-based functional connectivity between subprocesses and their relationship between psychopathologies. Our study also underscores the need for studying pediatric psychiatric disorders simultaneously rather than independently, as well as studying the within-disorder heterogeneity and the high rates of comorbidity among children with psychiatric disorders.

## Data Availability

The data that support the findings of this study are available upon reasonable request from the corresponding author.

## Acknowledgments

JRD is holder of a doctoral scholarship from the Fonds de Recherche du Québec en Santé (FRQS). SP is holder of the Eli Lilly Canada Chair on schizophrenia research.

We also would like to thank Dr. Angela Laird and the Neuroinformatics and Brain Connectivity Lab who have made publicly available the code that was used and modified for the specific purposes of the current study.

## Disclosures

SBE was supported by the Deutsche Forschungsgemeinschaft (DFG), the National Institute of Mental Health (R01-MH074457), the Helmholtz Portfolio Theme “Supercomputing and Modeling for the Human Brain”, ad the European Union’s Horizon 2020 Research and Innovation Programme under Grant Agreement No. 945539 (HBP SGA3)

## Notes

### Competing Interest Statement

The authors have declared no competing interest.

