## Supplementary Material for "Transdiagnostic Neural Signatures in common Pediatric Psychiatric Disorders: a data-driven meta-analysis of functional neuroimaging studies"

### Table of Contents

**Supplementary Table 1.** Included studies: Sample characteristics and fMRI tasks 3

**Supplementary Table 2.** Included studies: Medication and task-characteristics 6

**Supplementary Figure 1.** Metrics computed for K=2 to 15 clustering solutions 9

**Supplementary Figure 2.** Results from the forward (classical) ALE meta-analyses 10

**Supplementary Table 3.** Results from disorder-specific meta-analyses (p<0.001 unc.) 11

**Supplementary Table 4.** List of fMRI tasks per MAGs 12

**Supplementary Phenotype Assessment:** Literature Bias & Subanalyses 15

**Supplementary Table 5.** Disorder-specific sample and task characteristics 16

**Supplementary Table 6.** Disorder-specific comorbidities 16

**Supplementary Table 7.** Disorder-specific comorbidities per MAGs 17

**REFERENCES** 18

**Supplementary Table 1.** Included studies: Sample characteristics and fMRI task.

| Primary Diagnosis | First Author, Year | Experimental Task | TD (N=) | CASES (N=) | Male (%) | Mean Age |
|---|---|---|---|---|---|---|
| ADHD | Braet et al. 2011 (1) | Go-No/Go | 38 | 20 | 0.85 | 14.1 |
| ADHD | Bédard et al. 2014 (2) | N-Back | 21 | 24 | 0.88 | 12.4 |
| ADHD | Bollmann et al. 2017 (3) | Spatial Working Memory Task | 14 | 12 | 0.47 | 10.9 |
| ADHD | Booth et al. 2005 (4) | Go-No/Go | 12 | 12 | 0.67 | 11.0 |
| ADHD | Cao et al. 2008 (5) | Cued Target Detection | 13 | 12 | 1.00 | 13.4 |
| ADHD | Cerullo et al. 2009 (6) | Continuous Performance Task | 13 | 10 | 0.70 | 14.0 |
| ADHD | Chou et al. $2015_1$ (7) | Stroop Task | 20 | 25 | 0.85 | 10.5 |
| ADHD | Chou et al. $2015_2$ (7) | Stroop Task | 20 | 25 | 0.85 | 10.5 |
| ADHD | Christakou et al. 2013 (8) | Sustained Attention Task | 20 | 20 | 1.00 | 14.0 |
| ADHD | Durston et al. 2006 (9) | Go-No/Go | 11 | 11 | 1.00 | 13.9 |
| ADHD | Durston et al. $2007_1$ (10) | Go-No/Go | 12 | 12 | 1.00 | 14.9 |
| ADHD | Durston et al. $2007_2$ (10) | Go-No/Go | 12 | 12 | 1.00 | 14.9 |
| ADHD | Ercan et al. 2016 (11) | Go-No/Go | 100 | 50 | 0.58 | 11.0 |
| ADHD | Fan et al. 2014 (12) | Stroop Task | 23 | 25 | 0.92 | 10.9 |
| ADHD | Hart et al. 2014 (13) | Stop Task | 20 | 20 | 1.00 | 12.1 |
| ADHD | Hart et al. 2014 (14) | Stop Task | 30 | 30 | 1.00 | 13.9 |
| ADHD | Hauser et al. 2014 (15) | Reversal Learning Task | 20 | 20 | 0.65 | 14.6 |
| ADHD | Hwang et al. 2015 (16) | Stroop Paradigm | 35 | 26 | 0.65 | 14.5 |
| ADHD | Iannaccone et al. $2015_1$ (17) | Speeded Flanker Task | 18 | 18 | 0.61 | 14.5 |
| ADHD | Iannaccone et al. $2015_2$ (17) | Speeded Flanker Task | 18 | 18 | 0.61 | 14.5 |
| ADHD | Janssen et al. 2015 (18) | Stop-Signal Task | 17 | 21 | 0.91 | 10.6 |
| ADHD | Kohls et al. 2014 (19) | Rewarded Go-No/Go | 17 | 16 | 1.00 | 14.5 |
| ADHD | Konrad et al. 2006 (20) | Attention Network Test | 16 | 16 | 1.00 | 10.2 |
| ADHD | Krauel et al. 2007 (21) | Recognition Test | 12 | 12 | 1.00 | 14.5 |
| ADHD | Li et al. 2014 (22) | N-Back | 27 | 33 | 1.00 | 9.9 |
| ADHD | Ma et al. 2012 (23) | Go-No/Go | 15 | 15 | 0.53 | 9.9 |
| ADHD | Ma et al. 2016 (24) | Rewarded Stroop Task | 33 | 25 | 0.76 | 15.4 |
| ADHD | Massat et al. 2012 (25) | N-Back | 14 | 19 | 0.47 | 10.8 |
| ADHD | Massat et al. 2018 (26) | Stop Signal Task | 19 | 18 | 0.44 | 10.6 |
| ADHD | Metin et al. 2017 (27) | Spatial Attention Paradigm | 16 | 19 | 0.74 | 10.32 |
| ADHD | Mostofsky et al. 2006 (28) | Finger Sequencing Paradigm | 11 | 11 | 0.73 | 10.4 |
| ADHD | Norman et al. 2017 (29) | Sustained Attention Task | 20 | 20 | 1.00 | 15.0 |
| ADHD | Norman et al. 2018 (30) | Iowa Gambling Task | 20 | 16 | 1.00 | 14.6 |
| ADHD | O'Conaill et al. 2015 (31) | Visual Search Task | 21 | 19 | 0.90 | 11.9 |
| ADHD | Orinstein et al. 2014 (32) | Auditory Oddball Attention Task | 20 | 18 | 0.78 | 15.2 |
| ADHD | Passarotti et al. 2010 (33) | N-Back | 19 | 14 | 0.64 | 13.0 |
| ADHD | Passarotti et al. 2010 (34) | Emotional Valence Stroop Task | 14 | 15 | 0.80 | 12.9 |
| ADHD | Passarotti et al. 2010 (35) | Response Inhibition Task | 15 | 11 | 0.55 | 13.1 |
| ADHD | Poissant et al. 2016 (A) (36) | Pictures Story | 12 | 7 | 0.00 | 11.0 |
| ADHD | Poissant et al. 2016 (B) (36) | Pictures Story | 9 | 16 | 1.00 | 11.0 |
| ADHD | Posner et al. 2011 (37) | Subliminal Fearful Faces | 15 | 15 | 0.87 | 13.5 |
| ADHD | Posner et al. 2011 (38) | Emotional Stroop Task | 15 | 15 | 0.87 | 13.5 |
| ADHD | Rubia et al. 2005 (39) | Stop Task | 22 | 16 | 1.00 | 13.0 |
| ADHD | Rubia et al. 2008 (40) | Stop Task | 20 | 20 | 1.00 | 13.2 |
| ADHD | Rubia et al. 2009 (41) | Reward Continuous Performance | 16 | 18 | 1.00 | 13.3 |
| ADHD | Rubia et al. 2009 (42) | Simon Task | 20 | 20 | 1.00 | 13.2 |
| ADHD | Rubia et al. 2009 (43) | Delay Discounting Task | 10 | 10 | 1.00 | 14.0 |
| ADHD | Rubia et al. 2010 (44) | Visual–Spatial Switch Task | 20 | 14 | 1.00 | 13.3 |
| ADHD | Rubia et al. $2010_1$ (45) | Stop Task | 20 | 18 | 1.00 | 14.5 |

| | | | | | | |
|---|---|---|---|---|---|---|
| ADHD | Rubia et al. $2010_2$ (45) | Switch Task | - | 12 | 1.00 | 13.7 |
| ADHD | Rubia et al. 2011 (46) | Simon Task | 20 | 18 | 1.00 | 14.3 |
| ADHD | Schwarz et al. 2015 (47) | Anti-Saccades | 11 | 11 | 0.64 | 10.4 |
| ADHD | Sheridan et al. 2007 (48) | Match-to-Sample Task | 10 | 10 | 0.00 | 15.2 |
| ADHD | Silk et al. 2008 (49) | Raven's Progressive Matrices | 12 | 12 | 1.00 | 11.2 |
| ADHD | Siniatchkin et al. 2012 (50) | Go/NoGo | 27 | 17 | 0.83 | 9.3 |
| ADHD | Smith et al. 2008 (51) | Time Discrimination | 17 | 21 | 1.00 | 12.8 |
| ADHD | Spinelli et al. 2011 (52) | Go-No/Go | 17 | 13 | 0.69 | 10.6 |
| ADHD | Spinelli et al. 2011 (53) | Go-No/Go | 17 | 13 | 0.69 | 10.6 |
| ADHD | Stevens et al. $2017_1$ (A) (54) | Monetary Incentive Delay | 134 | 40 | 0.78 | 14.8 |
| ADHD | Stevens et al. $2017_2$ (A) (54) | Monetary Incentive Delay | - | - | 0.78 | 14.8 |
| ADHD | Stevens et al. $2017_1$ (B) (54) | Monetary Incentive Delay | - | 31 | 0.84 | 15.0 |
| ADHD | Stevens et al. $2017_2$ (B) (54) | Monetary Incentive Delay | - | - | 0.84 | 15.0 |
| ADHD | Stevens et al. 2017 (C) (54) | Monetary Incentive Delay | - | 46 | 0.80 | 15.1 |
| ADHD | Tamm et al. 2004 (55) | Go-No/Go | 12 | 10 | 1.00 | 16.0 |
| ADHD | Tamm et al. 2006 (56) | Oddball Task | 12 | 14 | 1.00 | 15.6 |
| ADHD | Tamm et al. 2012 (57) | Fluid Reasoning Task | 10 | 12 | 0.54 | 9.0 |
| ADHD | Tegelbeckers et al. 2015 (58) | Visual Oddball Task | 19 | 19 | 1.00 | 13.3 |
| ADHD | Vance et al. 2007 (59) | Mental Rotation Task | 12 | 12 | 1.00 | 11.1 |
| ADHD | van Ewijk et al. 2015 (60) | Spatial span task | 103 | 109 | 0.60 | 17.1 |
| ADHD | van Rooij et al. 2015 (61) | Stop-Signal Task | 111 | 185 | 0.70 | 17.3 |
| ADHD | Vetter et al. 2018 (62) | perceptual discrimination task | 25 | 25 | 1.00 | 14.3 |
| ADHD | Vloet et al. 2010 (63) | Spatial Stimulus–Response | 14 | 14 | 1.00 | 11.3 |
| ADHD | von Rhein et al. 2015 (64) | monetary incentive delay task | 108 | 150 | 0.70 | 17.7 |
| ADHD | Wang et al. 2013 (65) | Continuous Performance Task | 31 | 28 | 0.89 | 9.6 |
| ADHD | Hegarty et al. 2021 (66) | Stop-Signal Task | 30 | 30 | 0.67 | 10.1 |
| ADHD | Finger et al., 2008 (67) | Probabilistic Reversal Task | 14 | 14 | 0.71 | 13.4 |
| ADHD | Gatzke-Kopp et al. 2009 (68) | Monetary Incentive task | 11 | 19 | 1.00 | 13.6 |
| ADHD | Bubenzer-Busch et al. $2016_1$ (69) | Point Subtraction Aggression | 27 | 27 | 1.00 | 10.9 |
| ADHD | Bubenzer-Busch et al. $2016_2$ (69) | Point Subtraction Aggression | 27 | 27 | 1.00 | 10.9 |
| CD/ODD | Banich et al. 2007 (70) | Stroop Task | 12 | 12 | 1.00 | 16.7 |
| CD/ODD | Bjork et al. 2010 (71) | Monetary Incentive Delay Task | 12 | 12 | 1.00 | 15.4 |
| CD/ODD | Crowley et al. $2010_1$ (72) | Colorado Balloon Game | 20 | 20 | 1.00 | 16.5 |
| CD/ODD | Crowley et al. $2010_2$ (72) | Colorado Balloon Game | 20 | 20 | 1.00 | 16.5 |
| CD/ODD | Dong et al. 2017 (73) | Facial Expression (Pain) | 36 | 30 | 1.00 | 15.1 |
| CD/ODD | Fairchild et al. 2014 (74) | Emotional Face Task | 20 | 20 | 0.00 | 16.9 |
| CD/ODD | Fehlbaum et al. 2018 (75) | Affective Stroop Task | 39 | 39 | 0.74 | 15.9 |
| CD/ODD | Finger et al. $2011_1$ (76) | Probabilistic Reversal Task | 15 | 15 | 0.60 | 14.1 |
| CD/ODD | Finger et al. $2011_2$ (76) | Probabilistic Reversal Task | 15 | 15 | 0.60 | 14.1 |
| CD/ODD | Herpertz et al. 2008 (77) | Passive-Viewing (Emotional) | 22 | 22 | 1.00 | 14.7 |
| CD/ODD | Hwang et al. 2016 (A) (78) | Affective Stroop Task | 28 | 17 | 0.71 | 14.8 |
| CD/ODD | Hwang et al. 2016 (B) (78) | Affective Stroop Task | - | 18 | 0.56 | 14.6 |
| CD/ODD | Klapwijk et al., 2016a (79) | Empathic Emotional Face Task | 33 | 23 | 1.00 | 16.6 |
| CD/ODD | Klapwijk et al., 2016b (80) | Dictator Game | 33 | 32 | 1.00 | 16.8 |
| CD/ODD | Marsh et al., 2011 (81) | Implicit Association Test | 14 | 14 | 0.57 | 14.4 |
| CD/ODD | Marsh et al., 2013 (82) | Empathic Situation of Pain Task | 21 | 14 | 0.54 | 15.4 |
| CD/ODD | Passamonti et al., 2010 (A) (83) | Emotional Face Task | 40 | 27 | 1.00 | 17.7 |
| CD/ODD | Passamonti et al., 2010 (B) (83) | Emotional Face Task | - | 25 | 1.00 | 17.1 |
| CD/ODD | Rubia et al. 2008 (40) | Stop Task | 20 | 13 | 1.00 | 13.0 |
| CD/ODD | Rubia et al. 2009 (41) | Reward Continuous Performance | 16 | 14 | 1.00 | 12.8 |
| CD/ODD | Rubia et al. 2009 (42) | Simon Task | 20 | 13 | 1.00 | 12.9 |
| CD/ODD | Rubia et al. 2010 (44) | Visual–Spatial Switch Task | 20 | 14 | 1.00 | 12.6 |

| | | | | | | |
|---|---|---|---|---|---|---|
| CD/ODD | Schwenck et al., 2017 (84) | monetary gambling task | 24 | 19 | 1.00 | 13.8 |
| CD/ODD | Thornton et al., 2017 (85) | Animacy Attention Task | 20 | 29 | 0.69 | 14.7 |
| CD/ODD | White et al., 2012a (86) | Eye Gaze Task | 19 | 17 | 0.76 | 15.5 |
| CD/ODD | White et al., 2012b (87) | Emotion-Attention Bars task | 17 | 15 | 0.80 | 15.7 |
| CD/ODD | White et al., 2013$_{1}$ (88) | Passive Avoidance Task | 18 | 20 | 0.82 | 15.2 |
| CD/ODD | White et al., 2013$_{2}$ (88) | Passive Avoidance Task | 18 | 20 | 0.82 | 15.2 |
| CD/ODD | White et al., 2014 (89) | Doors Task | 15 | 15 | 0.73 | 14.4 |
| CD/ODD | Zhang et al., 2015 (90) | Go/Stop Task | 40 | 29 | 1.00 | 15.1 |
| CD/ODD | Zhu et al. 2014 (91) | Go/Stop Task | 10 | 11 | 1.00 | 11.5 |
| CD/ODD | Raschle et al. 2019 (92) | emotion-regulation task | 29 | 30 | 0.00 | 16.3 |
| ANX | Yang et al. 2004 (93) | Emotional Perception | 6 | 5 | 0.20 | 13.0 |
| ANX | Carrion et al. 2008 (94) | Go-No/Go | 14 | 16 | 0.56 | 13.7 |
| ANX | Keding-Herringa et al. 2016 (95) | Dynamic Face Task | 25 | 28 | 0.35 | 14.3 |
| ANX | Hart et al. 2018 (96) | emotion discrimination task | 27 | 20 | 0.70 | 17.5 |
| ANX | Carlisi et al. 2017 (97) | face-attention paradigm | 19 | 14 | 0.29 | 14.1 |
| ANX | Gold et al. 2020 (98) | threat conditioning | 47 | 53 | 0.45 | 12.8 |
| ANX | Thomas et al. 2001 (99) | Passive-Viewing Emotional Face | 12 | 12 | 0.58 | 12.8 |
| ANX | Monk et al. 2006 (100) | Emotional Face probe detection | 15 | 18 | 0.47 | 12.3 |
| ANX | Monk et al. 2008 (101) | Emotional Face visual probe task | 12 | 17 | 0.65 | 13.1 |
| ANX | Strawn et al. 2012 (102) | Emotional CPT | 10 | 10 | 0.40 | 14.3 |
| ANX | Yin et al. 2017 (103) | Valence Evaluation Task | 14 | 20 | 0.25 | 15.7 |
| ANX | Burkhouse et al. 2018 (104) | Emotional Conflict Task | 25 | 25 | 0.41 | 15.3 |
| ANX | Blair et al. 2011 (105) | Emotional Face Task | 16 | 14 | 0.50 | 13.3 |
| DEP | Chantiluke et al. 2012$_{1}$ (106) | Reward Continuous Performance | 21 | 20 | 0.50 | 16.2 |
| DEP | Chantiluke et al. 2012$_{2}$ (106) | Reward Continuous Performance | 21 | 20 | 0.50 | 16.2 |
| DEP | Colich et al. 2015 (107) | modified affective Go/No-Go | 15 | 18 | 0.17 | 15.6 |
| DEP | Davey et al. 2011 (108) | Being-Liked (Emotional Face) | 20 | 19 | 0.35 | 18.6 |
| DEP | Diler et al. 2013 (109) | emotional faces (gender) | 10 | 10 | 0.20 | 15.9 |
| DEP | Diler et al. 2014 (110) | Go/No-Go | 10 | 10 | 0.20 | 15.9 |
| DEP | Gaffrey et al. 2013 (111) | Facial Emotion Viewing Task | 31 | 23 | 0.57 | 5.0 |
| DEP | Halari et al. 2009$_{1}$ (112) | Simon Task | 21 | 21 | 0.48 | 16.2 |
| DEP | Halari et al. 2009$_{2}$ (112) | Switch Task | 21 | 21 | 0.48 | 16.2 |
| DEP | Halari et al. 2009$_{3}$ (112) | Stop Task | 21 | 21 | 0.48 | 16.2 |
| DEP | Hall et al. 2014 (113) | Emotional Faces Task | 23 | 32 | 0.26 | 15.5 |
| DEP | Roberson-Nay et al. 2006 (114) | Emotional Face Task: Encoding | 23 | 10 | 0.30 | 13.8 |
| DEP | Sharp et al. 2014 (115) | Modified card-guessing game | 19 | 14 | 0.00 | 13.4 |
| DEP | Tao et al. 2012 (116) | Emotional Face Task (gender) | 21 | 19 | 0.42 | 14.2 |
| DEP | Yang et al. 2009 (117) | Stop Task | 13 | 13 | 0.46 | 16.0 |
| DEP | Yang et al. 2010 (118) | facial-emotion matching task | 12 | 12 | 0.58 | 15.9 |
| DEP | Pan et al. 2011 (119) | Go/No-Go task | - | 15 | 0.47 | 15.9 |
| DEP | Pan et al. 2013 (A) (120) | Iowa Gambling Task | 13 | 15 | 0.27 | 16.2 |
| DEP | Pan et al. 2013 (B) (120) | Iowa Gambling Task | - | 14 | 0.50 | 15.8 |
| DEP | Groschwitz et al. 2016 (121) | Cyberball Task | 15 | 14 | 0.21 | 15.4 |
| DEP | Suzuki et al. 2014 (A) (122) | Emotional Face Task | 51 | 42 | 0.48 | 9.8 |
| DEP | Suzuki et al. 2014 (B) (122) | Emotional Face Task | - | 22 | 0.41 | 10.2 |
| DEP | De Bellis et Hooper 2013 (123) | Emotional oddball task | 5 | 5 | 0.40 | 15.5 |

*Note.* (A)(B) = distinct samples from the same study. $_{1\text{-}2}$ = distinct task contrasts or fMRI task from the same study. MED = Medication (in %); COG = Cognitive; RI = Response Inhibition; ATTN = Attention; EMO = Emotional; BOTH = Both Positive and Negative Emotion; POS = Positive; NEG = Negative;

Supplementary Table 2. Included studies: Medication and task-characteristics

| Primary Diagnosis | First Author, Year | MED (%) | Task-Characteristics | | | | | | | |
|---|---|---|---|---|---|---|---|---|---|---|
| | | | COG | RI | ATTN | EMO | BOTH | POS | NEG | Others |
| ADHD | Braet et al. 2011 (1) | NA | X | X | | | | | | |
| ADHD | Bédard et al. 2014 (2) | 0.08 | X | | | | | | | |
| ADHD | Bollmann et al. 2017 (3) | 0.67 | X | | | | | | | |
| ADHD | Booth et al. 2005 (4) | 1.00 | X | X | | | | | | |
| ADHD | Cao et al. 2008 (5) | 0.25 | X | | X | | | | | |
| ADHD | Cerullo et al. 2009 (6) | 0.00 | X | | X | | | | | |
| ADHD | Chou et al. 2015$_1$ (7) | 0.00 | X | X | | | | | | |
| ADHD | Chou et al. 2015$_2$ (7) | 0.00 | X | X | | | | | | |
| ADHD | Christakou et al. 2013 (8) | 0.00 | X | | X | | | | | |
| ADHD | Durston et al. 2006 (9) | 0.55 | X | X | | | | | | |
| ADHD | Durston et al. 2007$_1$ (10) | 0.75 | X | X | | | | | | |
| ADHD | Durston et al. 2007$_2$ (10) | 0.75 | X | X | | | | | | |
| ADHD | Ercan et al. 2016 (11) | NA | X | X | | | | | | |
| ADHD | Fan et al. 2014 (12) | 0.24 | X | X | | | | | | |
| ADHD | Hart et al. 2014 (13) | 0.30 | X | X | | | | | | |
| ADHD | Hart et al. 2014 (14) | 0.30 | X | X | | | | | | |
| ADHD | Hauser et al. 2014 (15) | 0.80 | | | | X | | X | | |
| ADHD | Hwang et al. 2015 (16) | 0.42 | X | X | | X | X | | | |
| ADHD | Iannaccone et al. 2015$_1$ (17) | 0.72 | X | X | | | | | | |
| ADHD | Iannaccone et al. 2015$_2$ (17) | 0.72 | X | X | | | | | | |
| ADHD | Janssen et al. 2015 (18) | 0.90 | X | X | | | | | | |
| ADHD | Kohls et al. 2014 (19) | 0.56 | | | | X | | X | | |
| ADHD | Konrad et al. 2006 (20) | 0.00 | X | | X | | | | | |
| ADHD | Krauel et al. 2007 (21) | 0.42 | X | | | | | | | |
| ADHD | Li et al. 2014 (22) | 0.00 | X | | | | | | | |
| ADHD | Ma et al. 2012 (23) | 0.20 | X | X | | | | | | |
| ADHD | Ma et al. 2016 (24) | 0.60 | | | | X | | X | | |
| ADHD | Massat et al. 2012 (25) | 0.00 | X | | | | | | | |
| ADHD | Massat et al. 2018 (26) | 0.00 | X | X | | | | | | |
| ADHD | Metin et al. 2017 (27) | 0.00 | | | | X | | X | | |
| ADHD | Mostofsky et al. 2006 (28) | 0.73 | | | | | | | | X |
| ADHD | Norman et al. 2017 (29) | 0.35 | X | | X | | | | | |
| ADHD | Norman et al. 2018 (30) | 0.50 | | | | X | X | | | |
| ADHD | O'Conaill et al. 2015 (31) | 0.79 | X | | X | | | | | |
| ADHD | Orinstein et al. 2014 (32) | NA | X | | X | | | | | |
| ADHD | Passarotti et al. 2010 (33) | 0.00 | | | | X | X | | | |
| ADHD | Passarotti et al. 2010 (34) | 0.00 | | | | X | X | | | |
| ADHD | Passarotti et al. 2010 (35) | 0.00 | X | X | | | | | | |
| ADHD | Poissant et al. 2016 (A) (36) | NA | X | | | | | | | |
| ADHD | Poissant et al. 2016 (B) (36) | NA | X | | | | | | | |
| ADHD | Posner et al. 2011 (37) | 0.00 | | | | X | | | X | |
| ADHD | Posner et al. 2011 (38) | 0.53 | | | | X | X | | | |
| ADHD | Rubia et al. 2005 (39) | 0.00 | X | X | | | | | | |
| ADHD | Rubia et al. 2008 (40) | 0.00 | X | X | | | | | | |
| ADHD | Rubia et al. 2009 (41) | 0.00 | X | | X | X | | X | | |
| ADHD | Rubia et al. 2009 (42) | 0.00 | X | X | | | | | | |
| ADHD | Rubia et al. 2009 (43) | 0.00 | | | | X | | X | | |
| ADHD | Rubia et al. 2010 (44) | 0.00 | X | | X | | | | | |

| | | | | | | | | | | |
|---|---|---|---|---|---|---|---|---|---|---|
| ADHD | Rubia et al. 2010$_1$ (45) | 0.00 | X | X | | | | | | |
| ADHD | Rubia et al. 2010$_2$ (45) | 0.00 | X | | X | | | | | |
| ADHD | Rubia et al. 2011 (46) | 0.00 | X | X | | | | | | |
| ADHD | Schwarz et al. 2015 (47) | 0.91 | | | | | | | | X |
| ADHD | Sheridan et al. 2007 (48) | 0.20 | X | | | | | | | |
| ADHD | Silk et al. 2008 (49) | 0.00 | X | | | | | | | |
| ADHD | Siniatchkin et al. 2012 (50) | 0.75 | X | X | | | | | | |
| ADHD | Smith et al. 2008 (51) | 0.00 | X | | | | | | | |
| ADHD | Spinelli et al. 2011 (52) | 0.15 | X | X | | | | | | |
| ADHD | Spinelli et al. 2011 (53) | 0.15 | X | X | | | | | | |
| ADHD | Stevens et al. 2017$_1$ (A) (54) | 0.68 | X | | | | | | | |
| ADHD | Stevens et al. 2017$_2$ (A) (54) | 0.68 | | | | X | | X | | |
| ADHD | Stevens et al. 2017$_1$ (B) (54) | 0.77 | X | X | | | | | | |
| ADHD | Stevens et al. 2017$_2$ (B) (54) | 0.77 | | | | X | | X | | |
| ADHD | Stevens et al. 2017 (C) (54) | 0.67 | X | | | | | | | |
| ADHD | Tamm et al. 2004 (55) | 0.50 | X | X | | | | | | |
| ADHD | Tamm et al. 2006 (56) | 0.36 | X | | X | | | | | |
| ADHD | Tamm et al. 2012 (57) | 0.00 | X | | | | | | | |
| ADHD | Tegelbeckers et al. 2015 (58) | 0.53 | X | | X | | | | | |
| ADHD | Vance et al. 2007 (59) | 0.00 | X | | | | | | | |
| ADHD | van Ewijk et al. 2015 (60) | 0.83 | X | | | | | | | |
| ADHD | van Rooij et al. 2015 (61) | 0.54 | X | X | | | | | | |
| ADHD | Vetter et al. 2018 (62) | NA | X | | | X | | | X | |
| ADHD | Vloet et al. 2010 (63) | NA | X | | X | | | | | |
| ADHD | von Rhein et al. 2015 (64) | 0.76 | | | | X | | X | | |
| ADHD | Wang et al. 2013 (65) | 0.00 | X | | X | | | | | |
| ADHD | Hegarty et al. 2021 (66) | 0.00 | X | X | | | | | | |
| ADHD | Finger et al., 2008 (67) | 0.71 | | | | X | X | | | |
| ADHD | Gatzke-Kopp et al. 2009 (68) | 0.53 | | | | X | | X | | |
| ADHD | Bubenzer-Busch et al. 2016$_1$ (69) | 0.85 | | | | X | | | X | |
| ADHD | Bubenzer-Busch et al. 2016$_2$ (69) | 0.85 | | | | X | | X | | |
| CD/ODD | Banich et al. 2007 (70) | NA | X | X | | | | | | |
| CD/ODD | Bjork et al. 2010 (71) | 0.67 | | | | X | | X | | |
| CD/ODD | Crowley et al. 2010$_1$ (72) | 0.30 | | | | X | | X | | |
| CD/ODD | Crowley et al. 2010$_2$ (72) | 0.30 | | | | X | | | X | |
| CD/ODD | Dong et al. 2017 (73) | NA | | | | X | | | X | |
| CD/ODD | Fairchild et al. 2014 (74) | NA | | | | X | | | X | |
| CD/ODD | Fehlbaum et al. 2018 (75) | 0.90 | X | X | | X | | | X | |
| CD/ODD | Finger et al. 2011$_1$ (76) | 0.47 | | | | X | | X | | |
| CD/ODD | Finger et al. 2011$_2$ (76) | 0.47 | | | | X | | | X | |
| CD/ODD | Herpertz et al. 2008 (77) | NA | | | | X | | | X | |
| CD/ODD | Hwang et al. 2016 (A) (78) | 0.24 | X | X | | X | | | X | |
| CD/ODD | Hwang et al. 2016 (B) (78) | 0.28 | X | X | | X | | | X | |
| CD/ODD | Klapwijk et al., 2016a (79) | 0.00 | | | | X | | | X | |
| CD/ODD | Klapwijk et al., 2016b (80) | 0.00 | X | | | | | | | |
| CD/ODD | Marsh et al., 2011 (81) | 0.43 | X | | | | | | | |
| CD/ODD | Marsh et al., 2013 (82) | 0.14 | X | | | X | | | X | |
| CD/ODD | Passamonti et al., 2010 (A) (83) | NA | | | | X | | | X | |
| CD/ODD | Passamonti et al., 2010 (B) (83) | NA | | | | X | | | X | |
| CD/ODD | Rubia et al. 2008 (40) | 0.00 | X | X | | | | | | |
| CD/ODD | Rubia et al. 2009 (41) | 0.00 | X | | X | X | X | | | |
| CD/ODD | Rubia et al. 2009 (42) | 0.00 | X | X | | | | | | |

| | | | | | | | | | |
|---|---|---|---|---|---|---|---|---|---|
| CD/ODD | Rubia et al. 2010 (44) | 0.00 | X | | X | | | | |
| CD/ODD | Schwenck et al., 2017 (84) | 0.33 | | | | X | | | X |
| CD/ODD | Thornton et al., 2017 (85) | 0.14 | X | | X | | | | |
| CD/ODD | White et al., 2012a (86) | 0.12 | X | | X | X | | | X |
| CD/ODD | White et al., 2012b (87) | 0.12 | | | | X | | | X |
| CD/ODD | White et al., 2013$_1$ (88) | 0.10 | | | | X | | X | |
| CD/ODD | White et al., 2013$_2$ (88) | 0.10 | | | | X | | | X |
| CD/ODD | White et al., 2014 (89) | 0.20 | | | | X | X | | |
| CD/ODD | Zhang et al., 2015 (90) | 0.00 | X | X | | | | | |
| CD/ODD | Zhu et al. 2014 (91) | 0.00 | X | X | | | | | |
| CD/ODD | Raschle et al. 2019 (92) | NA | | | | X | | | X |
| ANX | Yang et al. 2004 (93) | 0.00 | | | | X | | | X |
| ANX | Carrion et al. 2008 (94) | 0.00 | X | X | | | | | |
| ANX | Keding-Herringa et al. 2016 (95) | 0.00 | | | | X | X | | |
| ANX | Hart et al. 2018 (96) | 0.00 | | | | X | | | X |
| ANX | Carlisi et al. 2017 (97) | 0.00 | | | | X | | | X |
| ANX | Gold et al. 2020 (98) | 0.00 | | | | X | | | X |
| ANX | Thomas et al. 2001 (99) | 0.00 | | | | X | | | X |
| ANX | Monk et al. 2006 (100) | 0.00 | | | | X | | | X |
| ANX | Monk et al. 2008 (101) | 0.00 | | | | X | | | X |
| ANX | Strawn et al. 2012 (102) | 0.00 | X | | X | X | X | | |
| ANX | Yin et al. 2017 (103) | 0.00 | | | | X | | | X |
| ANX | Burkhouse et al. 2018 (104) | 0.00 | | | | X | | | X |
| ANX | Blair et al. 2011 (105) | 0.00 | | | | X | | | X |
| DEP | Chantiluke et al. 2012$_1$ (106) | 0.00 | X | | X | X | | X | |
| DEP | Chantiluke et al. 2012$_2$ (106) | 0.00 | X | | X | X | | X | |
| DEP | Colich et al. 2015 (107) | 0.39 | X | X | | X | | | X |
| DEP | Davey et al. 2011 (108) | 0.47 | | | | X | | | |
| DEP | Diler et al. 2013 (109) | 0.60 | | | | X | X | | |
| DEP | Diler et al. 2014 (110) | 0.47 | X | X | | | | | |
| DEP | Gaffrey et al. 2013 (111) | 0.00 | | | | X | X | | |
| DEP | Halari et al. 2009$_1$ (112) | 0.00 | X | X | | | | | |
| DEP | Halari et al. 2009$_2$ (112) | 0.00 | X | | X | | | | |
| DEP | Halari et al. 2009$_3$ (112) | 0.00 | X | X | | | | | |
| DEP | Hall et al. 2014 (113) | 0.00 | | | | X | | | X |
| DEP | Roberson-Nay et al. 2006 (114) | 0.00 | X | | | X | X | | |
| DEP | Sharp et al. 2014 (115) | 0.00 | | | | X | | X | |
| DEP | Tao et al. 2012 (116) | 0.00 | | | | X | | | X |
| DEP | Yang et al. 2009 (117) | 0.00 | X | X | | | | | |
| DEP | Yang et al. 2010 (118) | 0.00 | | | | X | X | | |
| DEP | Pan et al. 2011 (119) | 0.47 | X | X | | | | | |
| DEP | Pan et al. 2013 (A) (120) | 0.67 | | | | X | X | | |
| DEP | Pan et al. 2013 (B) (120) | 0.43 | | | | X | X | | |
| DEP | Groschwitz et al. 2016 (121) | 0.36 | | | | X | | | X |
| DEP | Suzuki et al. 2014 (A) (122) | NA | | | | X | | | X |
| DEP | Suzuki et al. 2014 (B) (122) | NA | | | | X | | | X |
| DEP | De Bellis et Hooper 2013 (123) | 0.00 | X | | X | X | | | X |

*Note.* (A)(B) = distinct samples from the same study. $_{1\text{-}2}$ = distinct task contrasts or fMRI task from the same study. MED = Medication (in %); COG = Cognitive; RI = Response Inhibition; ATTN = Attention; EMO = Emotional; BOTH = Both Positive and Negative Emotion; POS = Positive; NEG = Negative;

**Supplementary Figure 1.** Metrics computed for K=2 to 15 clustering solutions. **A.** This line graph represents silhouette (Rousseeuw, 1986) and calinski-harabasz (Calinski et Harabasz, 1974) metrics averaged over 5,000 subsampling iterations and then normalized against a null distribution. Green line = Silhouette metric, Blue line = Calinski-Harabasz metric. **B.** Line graph refers to changes in silhouette and calinski-harabasz values from K to K+1 (i.e. (K+1) – 1). Silhouette metrics (green) show a stable solution at k=8, while calinski-harabasz metric was monotonic. **C.** Adjusted Rand Indices between $K_{TRUE}$ - $K_{NULL}$. This plot shows lowest scores at K=3, K=8. **D.** Variation of Information between $K_{TRUE}$ - $K_{NULL}$. Although scores are rather monotonic, greatest increases moving from K=2 to 3, K=6 to K=7 and K=7 to K=8.

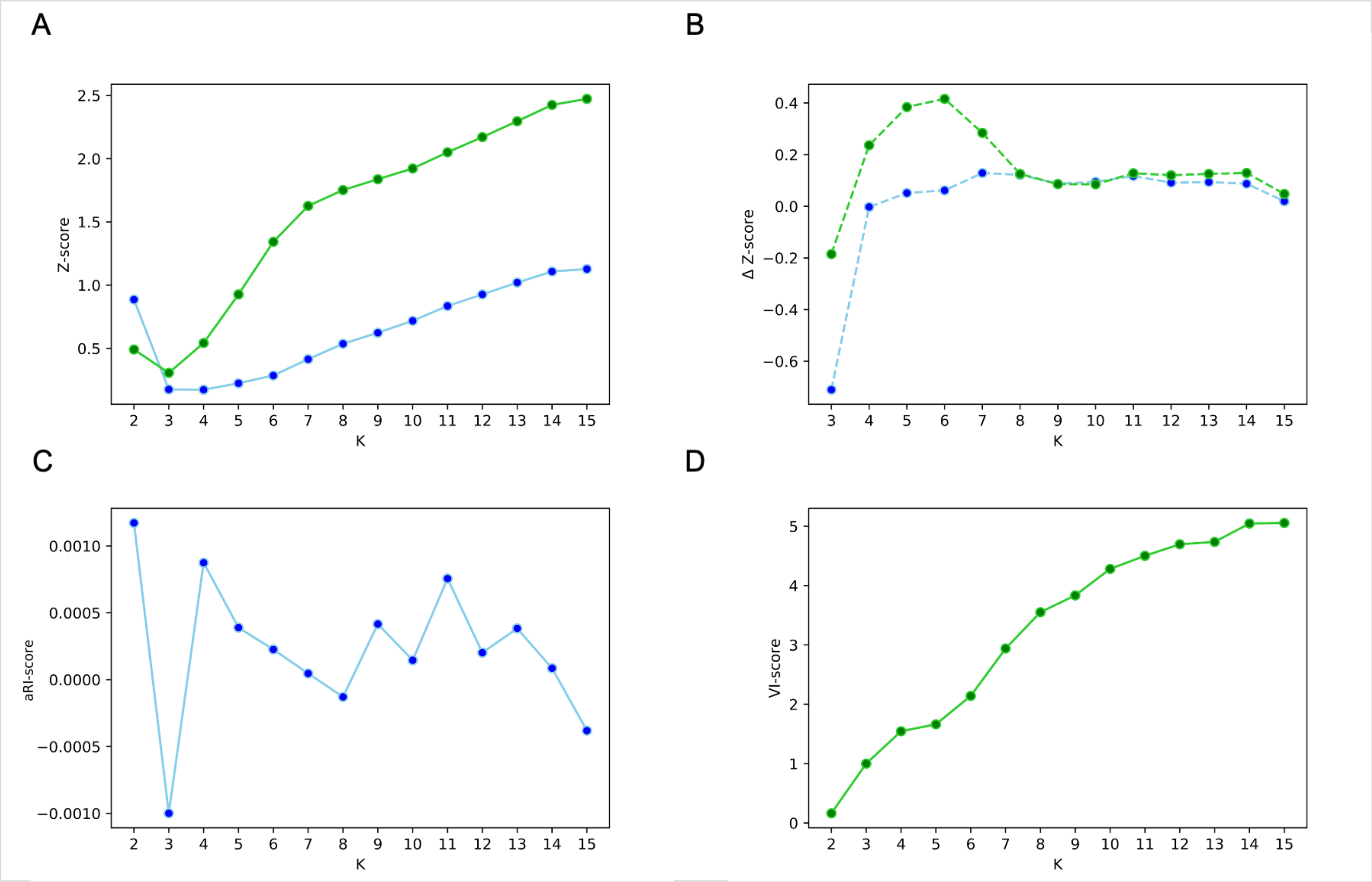

**Supplementary Figure 2.** Results from the forward (classical) ALE meta-analysis. Red cluster = Transdiagnostic Neurobiological Marker when pooling ADHD, CD/ODD, ANX and ADHD (x=4, y=24, z=42, ALE score=0.021, 323 voxels). Green Cluster = Externalizing Disorders (x=4, y=26, z=44, maximum ALE score=0.0279, 147 voxels); Blue Cluster = Internalizing Disorders (x=-8, y=38, z=8, maximum ALE score = 0.0167, 129 voxels).

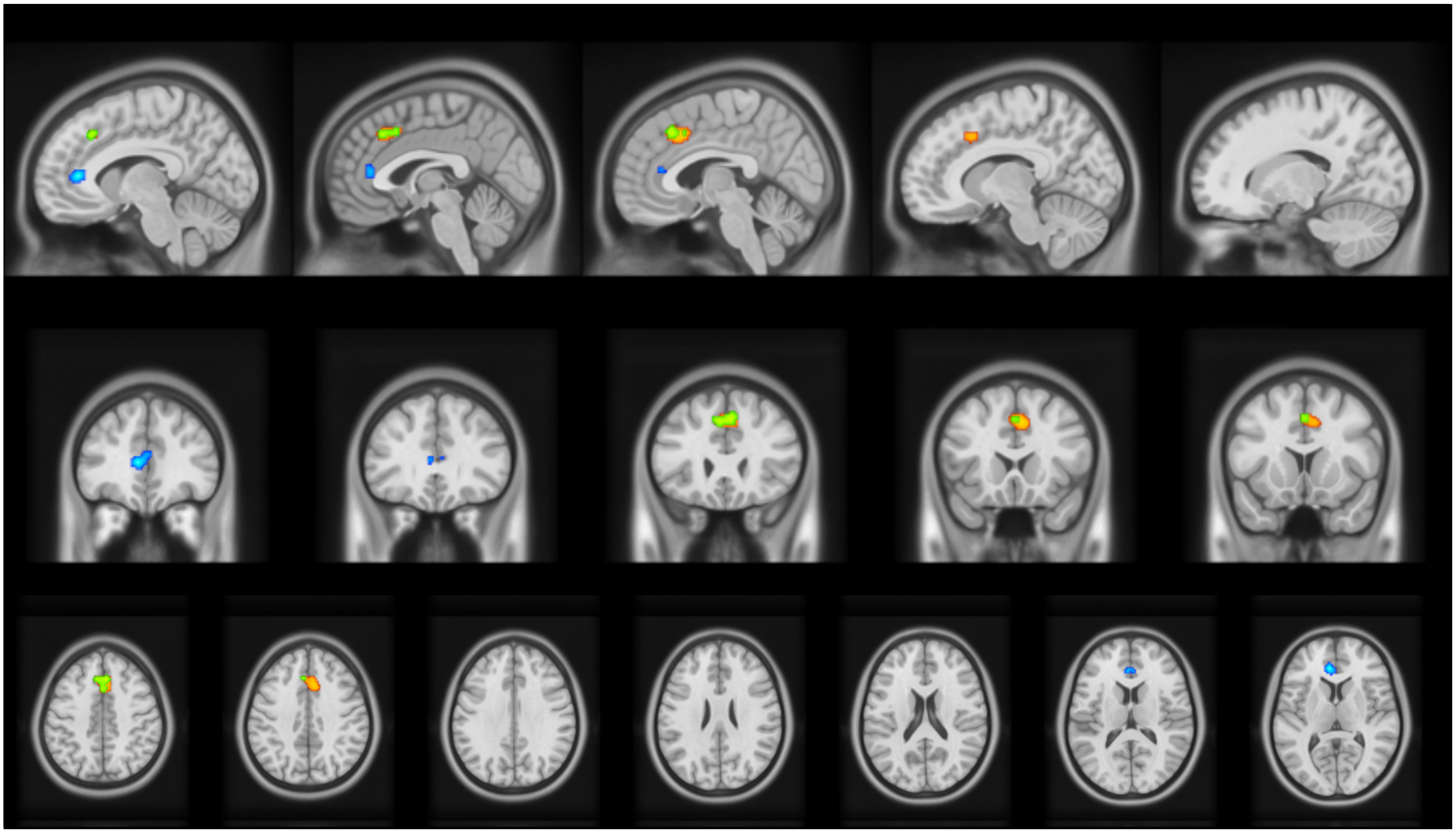

**Supplementary Table 3.** Results from classical meta-analysis on nosological categories

| Diagnosis Category | MNI Coordinates | | | ALE | Cluster Size |
|---|---|---|---|---|---|
| | X | Y | Z | | |
| *ADHD* | | | | | |
| dACC | -2 | 12 | 22 | 0.0198 | 51 |
| vlPFC (BA47) | 50 | 14 | -4 | 0.0179 | 35 |
| *CD/ODD* | | | | | |
| Caudate | 12 | 8 | 0 | 0.0118 | 40 |
| pre-SMA | -6 | 26 | 42 | 0.0113 | 28 |
| *ANX* | | | | | |
| N.S. | - | - | - | - | - |
| *DEP* | | | | | |
| N.S. | - | - | - | - | - |

*Note.* These results were performed using liberal threshold (p=0.001 uncorrected at a voxel-level). ADHD = Attention deficit hyperactivity disorder; CD/ODD = Conduct Disorder / Oppositional Defiant Disorder; ANX = Anxiety Disorder; DEP = Depressive Disorder. MNI = Montreal Neurological Institute; N.S. = Not Statistically Significant. dACC = dorsal Anterior Cingulate Cortex; vlPFC = ventrolateral Prefrontal Cortex; pre-SMA = pre-Supplementary Motor Area.

**Supplementary Table 4.** List of fMRI tasks per MAGs.

| **MAG1** |
|---|
| N-Back |
| Go-No/Go |
| Go-No/Go |
| Reversal Learning Task |
| Rewarded Go-No/Go |
| Rewarded Stroop Task |
| Response Inhibition Task |
| Stop Task |
| Simon Task |
| Go-No/Go |
| Spatial span task |
| monetary incentive delay task |
| Emotional face probe detection task |
| Continuous Performance Task with emotional distractors |
| face-attention paradigm |
| Colorado Balloon Game |
| Colorado Balloon Game |
| Probabilistic Reversal Task |
| Monetary Incentive task |
| Simon Task |
| Probabilistic Reversal Task |

| **MAG2** |
|---|
| Go-No/Go |
| Spatial Working Memory Task |
| Go-No/Go |
| Cued Target Detection |
| Stroop Task |
| Continuous Performance Task |
| Sustained Attention Task |
| Go-No/Go |
| Stroop Task |
| Stop Task |
| Speeded Flanker Task |
| Speeded Flanker Task |
| Stop-Signal Task |
| Attention Network Test |
| Recognition Test |
| N-Back |
| N-Back |
| Spatial Attention Paradigm |
| Sustained Attention Task |
| Iowa Gambling Task |
| Visual Search Task |
| Auditory Oddball Attention Task |
| N-Back |
| Emotional Valence Stroop Task |
| Subliminal Presentation of Fearful Faces |
| Emotional Stroop Task |
| Visual–Spatial Switch Task |
| Stop Task |
| Switch Task |
| Raven’s Progressive Matrices Task |
| Go/NoGo |
| Time Discrimination |
| Monetary incentive delay task |
| Monetary incentive delay task |
| Monetary incentive delay task |

monetary incentive delay task
Go-No/Go
Stop-Signal Task
Continuous Performance Task
Passive-Viewing Emotional Face Task
Emotional Face visual probe task
Valence Evaluation Task
Emotional Conflict Task
Emotional Face Task
Reward Continuous Performance Task
modified affective Go/No-Go
Being-Liked Task (Emotional Face)
emotional facial expression gender labeling task
Go/No-Go
Simon Task
Stop Task
Emotional Faces Task
Emotional Face Task: Encoding
Modified card-guessing game (monetary reward)
Emotional Face Task (gender labelling)
Parametric inhibitory task (Stop task)
facial-emotion matching task
Go/No-Go task
Iowa Gambling Task
Iowa Gambling Task
Emotional Perception
Go-No/Go
Dynamic Face Task
Emotional oddball task
Emotional oddball task
Cyberball Task
Stop-Signal Task
GoStop Task
Stroop Task
Monetary Incentive Delay (MID) task
Point Subtraction Aggression Game
Point Subtraction Aggression Game
Affective Stroop Task
Passive-Viewing Emotional Stimuli Task
Affective Stroop Task
Affective Stroop Task
Empathic Emotional Face Task
Dictator Game
Empathic Situation of Pain Task
Emotional Face Task
Emotional Face Task
Monetary gambling task
Animacy Attention Task
Passive Avoidance Task
Passive Avoidance Task
GoStop Task
Stroop Task

**MAG3**

N-Back
Finger Sequencing Paradigm
Stop Task
Simon Task
Anti-Saccade and Fixation Trials
Match-to-Sample Task
Oddball Task
Mental Rotation Task

Facial Emotion Viewing Task
Stop Task
Visual–Spatial Switch Task
Eye Gaze Task
Doors Task

**MAG4**

Go-No/Go
Stop Task
Stroop Paradigm
Go-No/Go
Pictures Story
Reward Continuous Performance Task
Fluid Reasoning Task
Visual Oddball Task
Threat conditioning
Emotional Face Task
Implicit Association Test (words)
Rewarded Continuous Performance Test (CPT)

*Supplementary Phenotype Assessment: Literature Bias & Subanalyses*

We tested if fMRI literature showed differences between nosological categories regarding the choice of neurocognitive task domains, average of sex ratio, and the average of prescribed medication per samples. Literature showed significant differences in cognitive tasks ($X_2$=23.75, $p<0.001$), general emotional tasks ($X_2$=42.11, $p<0.001$) but more specifically in tasks using simultaneously positive and negative stimuli ($X_2$=7.85, p=0.049) and negative stimuli only ($X_2$=53.69, $p<0.001$). Biases were also observed in the average of sex ratio ($X_2$=64.69, $p<0.001$) and the average of sample with prescribed medication ($X_2$=18.02, $p<0.001$).

Given that these factors may alter the relationships between nosological categories and MAGs, further analyses were performed by restricting experiments using specific cognitive task contrasts, emotional task contrasts, medication naïve and mixed sex samples. Subanalyses on emotion-specific stimuli (e.g., positive, negative or both) could not be performed due to the limited sample size across MAGs. First, the lower rates of DEP samples across MAG1 compared to other MAGs was only replicated when restricting experiments to those using an emotional task contrast ($X_2$=4.34, p=0.037) and a mixed sex samples ($X_2$=3.89, p=0.049). Furthermore, MAG1 had higher rates of ADHD samples in experiments with an emotional task contrast ($X_2$=3.18, p=0.050).

Finally, the relationship between MAG2 and DEP samples was replicated when restricting experiments to those using a cognitive task contrast ($X_2$=5.53, p=0.019), an emotional task contrast ($X_2$=5.22, p=0.022), but also in experiments with only medication naïve sample ($X_2$=5.70, p=0.017) and in mixed sex samples ($X_2$=7.54, p=0.006).

**Supplementary Table 5.** Disorder-specific sample and task characteristics

| Characteristics | ADHD | CD/ODD | ANX | DEP |
|---|---|---|---|---|
| *Task-domain* | | | | |
| Cognitive | 60 (75.9%) | 15 (46.9%) | 2 (14.3%) | 11 (50.0%) |
| RI | 29 (36.7%) | 8 (25%) | 1 (7.1%) | 6 (27.3%) |
| Attention | 14 (17.7%) | 4 (12.5%) | 1 (7.1%) | 4 (18.2%0 |
| Emotion | 19 (24.1%) | 23 (71.9%) | 13 (92.9%) | 16 (72.7%) |
| Positive | 10 (12.7%) | 4 (12.5%) | 0 (0%) | 3 (13.6%) |
| Negative | 3 (3.8%) | 17 (53.1%) | 11 (78.6%) | 6 (27.3%) |
| Both | 6 (7.6%) | 2 (6.3%) | 2 (14.3%) | 6 (27.3%) |
| *Sample* | | | | |
| Average Med per sample (Mean, SD) | 35.5 (33.9) | 21.5 (23.7) | 0 | 19.26 (25.0) |
| Average Boys per Sample (Mean, SD) | 82.2 (21.4) | 81.4 (27.3) | 44.4 (14.7) | 37.5 (15.2) |

Note. RI = Response Inhibition; Both = Both Positive and Negative Emotional Stimuli; Med = Medication

**Supplementary Table 6.** Disorder-specific comorbidities

| | Comorbidities | | | | | | | |
|---|---|---|---|---|---|---|---|---|
| | ADHD | | CD/ODD | | ANX | | DEP | |
| Primary Diagnosis | n | mean % | n | mean % | n | mean % | n | mean % |
| ADHD (n=79) | - | - | 65 | 14.4 (16.3) | 64 | 1.7 (4.6) | 65 | 1.2 (4.7) |
| CD/ODD (n=32) | 29 | 33,4 (24.1) | - | - | 31 | 1.0 (3.1) | 31 | 5.9 (19.6) |
| ANX (n=14) | 6 | 10.5 (15.6) | 10 | 6.7 (14.9) | - | - | 12 | 12 (28.2) |
| DEP (n=22) | 20 | 7.2 (9.0) | 20 | 4.1 (8.8) | 21 | 23.6 (27.1) | - | - |

Note. *n* = the number of samples with available data.

**Supplementary Table 7.** Disorder-specific comorbidities per MAGs

| Characteristics | MAG1 (k=21) | | MAG2 (k=87) | | MAG3 (k=13) | | MAG4 (k=12) | |
|---|---|---|---|---|---|---|---|---|
| | n | % | n | % | n | % | n | % |
| ***ADHD samples*** | | | | | | | | |
| ANX | 12 | 3.33% | 32 | 1.53% | 8 | 1.13% | 7 | 1.14% |
| DEP | 11 | 3.46% | 34 | 0.65% | 8 | 2.25% | 7 | 0% |
| CD/ODD | 12 | 23.58% | 33 | 15.50% | 8 | 6.63% | 7 | 5.14% |
| ***ANX samples*** | | | | | | | | |
| ADHD | 0 | 0% | 4 | 4.50% | 0 | 0% | 1 | 4% |
| DEP | 2 | 46.50% | 8 | 18.13% | 0 | 0% | 1 | 0% |
| CD/ODD | 2 | 0% | 6 | 2.50% | 0 | 0% | 1 | 4% |
| **DEP samples** | | | | | | | | |
| ADHD | 0 | 0% | 16 | 6.75% | 1 | 13% | 0 | 0% |
| ANX | 0 | 0% | 18 | 24.50% | 1 | 30% | 0 | 0% |
| CD/ODD | 0 | 0% | 16 | 2.06% | 1 | 17% | 0 | 0% |
| **CD/ODD samples** | | | | | | | | |
| ADHD | 2 | 33.50% | 16 | 36.19% | 4 | 25% | 3 | 26.30% |
| ANX | 4 | 0% | 16 | 1.25% | 4 | 0% | 3 | 1.67% |
| DEP | 4 | 0% | 16 | 1.69% | 4 | 0% | 3 | 15% |

*Note. n* = the number of samples with available data. The only analyses were performed in samples with ADHD and revealed marginally significant differences between MAGs relatively to comorbidities with CD/ODD (H=7.11, p=0.068). Post-hoc tests suggested that in ADHD samples, rate of CD/ODD comorbidity was higher is MAG1 compared to MAG3 (p=0.012) and MAG4 (p=0.011) but not compared to MAG2 (p=0.142). No differences were observed for ANX or DEP comorbidities in ADHD samples
